# Smoking-dependent expression alterations in nasal epithelium reveal immune impairment linked to germline variation and lung cancer risk

**DOI:** 10.1101/2021.11.24.21266740

**Authors:** Maria Stella de Biase, Florian Massip, Tzu-Ting Wei, Federico M. Giorgi, Rory Stark, Amanda Stone, Amy Gladwell, MarJn O’Reilly, Daniel Schütte, Ines de Santiago, Kerstin B. Meyer, Florian Markowetz, Bruce A J Ponder, Robert C Rintoul, Roland F Schwarz

## Abstract

Lung cancer is the leading cause of cancer-related death in the world. In contrast to many other cancers, a direct connection to modifiable lifestyle risk in the form of tobacco smoke has long been established. More than 50% of all smoking-related lung cancers occur in former smokers, often many years after smoking cessation. Despite extensive research, the molecular processes for persistent lung cancer risk are unclear.

To examine whether risk stratification in the clinic and in the general population can be improved upon by the addition of genetic data, and to explore the mechanisms of the persisting risk in former smokers, we have analysed transcriptomic data from accessible airway tissues of 487 subjects, including healthy volunteers and clinic patients of different smoking status. We developed a model to assess smoking associated gene expression changes and their reversibility after smoking is stopped, comparing healthy subjects to clinic patients with and without lung cancer. We find persistent smoking-associated immune alterations to be a hallmark of the clinic patients. Integrating previous GWAS data using a transcriptional network approach, we demonstrate that the same immune and interferon related pathways are strongly enriched for genes linked to known genetic risk factors, demonstrating a causal relationship between immune alteration and lung cancer risk. Finally, we used accessible airway transcriptomic data to derive a non-invasive lung cancer risk classifier.

Our results provide initial evidence for germline-mediated personalised smoke injury response and risk in the general population, with potential implications for managing long-term lung cancer incidence and mortality.

## Introduction

Through international efforts and public health campaigns the prevalence of cigarette smoking worldwide has substantially decreased during the last 30 years (*1*). However, lung cancer remains a major cause of death in current and former smokers: over 40% of all lung cancers occur more than 15 years after smoking cessation (*2, 3*). Low-dose CT screening studies in asymptomatic smokers and former smokers, stratified for risk by age and smoking history, have shown a reduction in lung cancer related death by up to 26% (*4, 5*). Although CT lung screening has been demonstrated to be cost-effective (*6, 7*), improvements in risk stratification of participants could further improve cost-effectiveness thereby making screening more widely accessible and allowing detection of at-risk subjects overlooked by the current criteria.

Transcriptional profiles from normal airway epithelium have been proposed as potential molecular biomarkers of a personalised smoke-injury response related to increased risk, and as potential predictors of the presence of lung cancer. Early studies of bronchial cells provided a broad characterisation of the genes affected by cigarette smoke exposure (*8*) and their post-cessation reversibility (*9*), and included initial attempts to derive predictive cancer gene expression signatures (*10*). Following the model of a ‘field of injury’ throughout the airway epithelium, later efforts focused on more accessible tissues from the nasal or buccal cavity to assess the personal smoke injury response (*11, 12*). Sridhar et al. (*13*) and Zhang et al. (*14*) provided initial evidence on 25 patients that nasal epithelium might act as a proxy for smoking-induced gene expression changes in the bronchus. More recently, the AEGIS study team presented a large multi-centre study in which they showed that a classifier based on microarray gene expression data in bronchial epithelium improved the diagnostic performance of bronchoscopy in patients being investigated for suspected lung cancer (*15*). They followed this up with a similar study based on nasal gene expression (*16*). They showed significant concordance between gene expression in bronchial and nasal epithelium, and that a lung cancer classifier based on nasal gene expression together with clinical risk factors had significantly improved predictive performance over a classifier based on clinical risk factors alone. These studies addressed the question of improving the diagnostic management of current and former smokers in whom lung cancer is already suspected due to the presence of pulmonary nodules detected during CT screening.

To date, no study so far addresses the important question of whether and how the smoke-injury response differs in the general population from that observed in individuals with an elevated prescreening risk. Accordingly, no molecular risk stratification strategy exists for the general population, where any early detection measures would arguably reap the greatest benefits. Here, we present a cohort which includes current and former smokers with suspected lung cancer based on clinical evaluation from a physician, as well as a group of never, former and current smoker healthy volunteers from the general population (Fig. 1). Our study provides an in-depth characterisation of the smoke-injury gene expression response in the healthy volunteers, based on accessible nasal tissue, and investigates the differences in smoke injury response between the healthy volunteers and the group of patients referred to the clinic. We derive molecular classifiers for assessing cancer risk in the clinic population as well as for predicting risk among the general population of asymptomatic current and former smokers. Using germline genotype data we associate individual differences in smoke injury response with known lung cancer GWAS risk loci, providing strong evidence for causal involvement of inherited variation in immune and interferon-related pathways, and for a role of immunosuppression in lung cancer development (*17, 18*).

**Figure 1:**
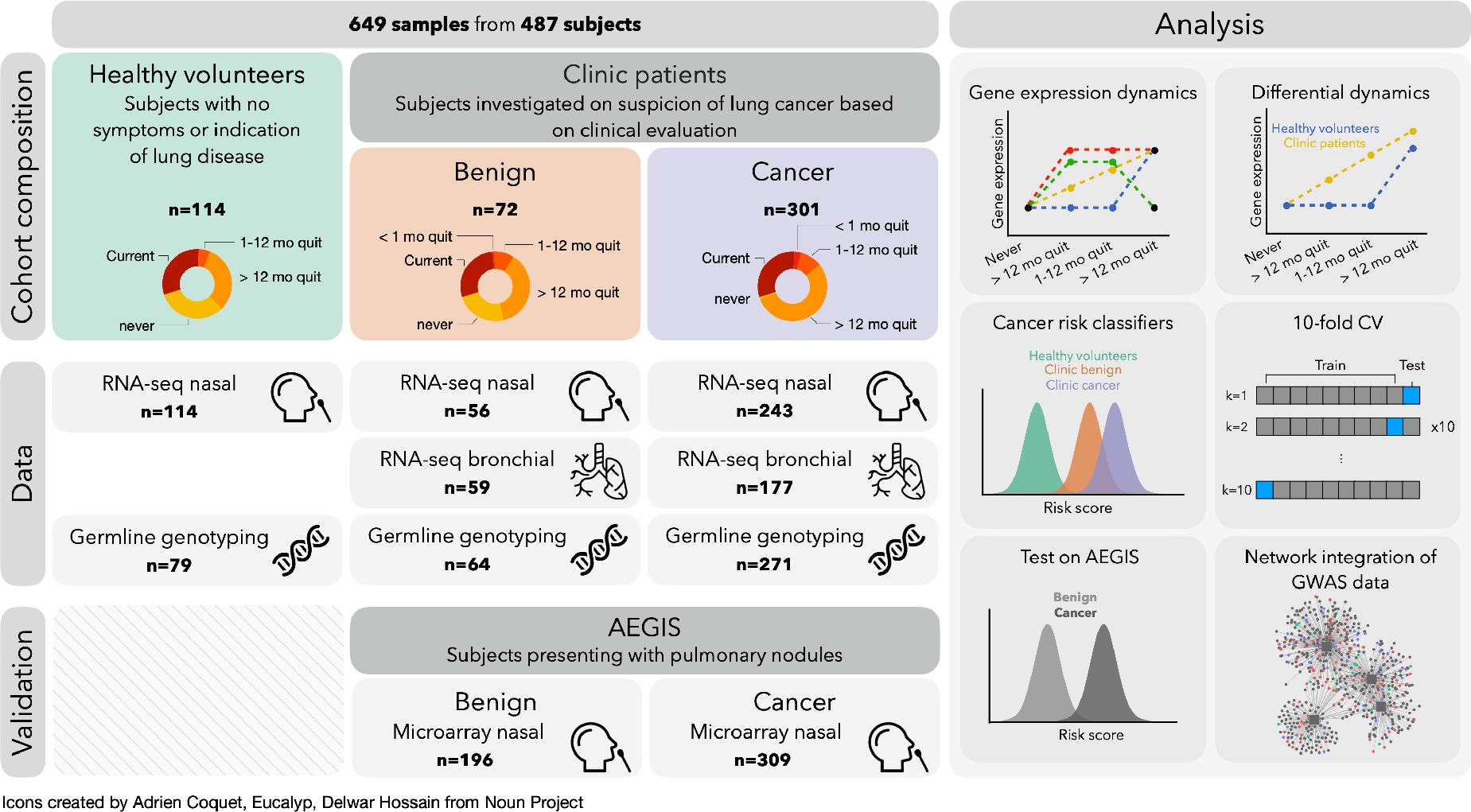
Overview of study subjects and data analysis. **(LeG)** Repartition of the subjects into clinical categories and smoking status. For each category, we show the number of subjects for which RNA-seq (on nasal and bronchial samples) and array-based blood genotyping were performed. Nasal samples from the AEGIS cohort were used as a validation set. **(Right)** Schematic of the different analyses conducted to stratify patients and identify dysregulated pathways among clinic patients.

## Results

### Study subjects

We recruited 487 subjects among which were 114 healthy volunteers from the Cambridge Bioresource (https://www.cambridgebioresource.group.cam.ac.uk/) and 373 patients referred to the outpatient clinic at Royal Papworth Hospital (Cambridge, UK) or Peterborough City Hospital (Peterborough, UK) with symptoms or imaging suspicious for lung cancer (clinic group). Healthy volunteers are here defined as individuals without any prior history or current suspicion for airway malignancy. Within the clinic group, 301 patients were diagnosed with cancer and 72 patients had a final diagnosis of a benign condition, the majority of which were inflammatory or infectious diseases (Fig. 1, Sup. Tab. 1). From these donors we collected a total of 649 samples: 413 nasal epithelial samples by mini-curette from 114 healthy donors and 299 clinic patients, and 236 bronchial brushings from clinic patients (Fig. 1, Methods). For 162 clinic patients both nasal and bronchial samples were collected (Sup. Tab. 2). Samples from healthy volunteers and clinic patients were collected and processed by the same staff using identical experimental protocols.

Smoking history was obtained for all subjects, confirmed by cotinine test, and recorded as never smokers (NV, n=45), current smokers (CS, n=153) and former smokers (FS, n=289). Former smokers were stratified into 3 categories based on their time from smoking cessation: former smokers who had quit less than one month (n=10), 1 to 12 months (n=45), or more than 1 year (n=234, median=168 months) prior to sample collection (Fig. 1, Methods). Cumulative smoke exposure was measured in pack-years, and stratified into 4 categories: none, 0-10, 11-30, >31 pack-years. In addition to smoking status, sex, age, lung cancer subtype and stage and presence of chronic obstructive pulmonary disease (COPD) were recorded according to the GOLD criteria (Vogelmeier et al. 2017) (Sup. Tab. 2). While most clinic patients with cancer were diagnosed with non-small cell lung cancer (NSCLC; n=245), 56 subjects presented with metastatic disease from an extra-thoracic primary (n=8), small-cell lung cancer (SCLC, n=31), or a rare pulmonary cancer e.g. carcinoid (n=17). Given the different underlying biology between NSCLC and other types of tumours, these subjects (with cancer status marked as *Ineligible* in Sup. Tab. 2) were included in all analyses investigating smoke injury response, but were excluded for lung cancer risk prediction. Clinic patients with a final diagnosis of a benign condition were followed up for a minimum of 1 year to confirm absence of cancer.

Airway samples underwent RNA sequencing using standard protocols. Blood samples were taken from 467 subjects for germline genotyping with Illumina Infinium Oncoarray pla{orm at 450K tagging germline variants. Total gene expression was quantified as variance-stabilised counts and corrected for batch effects in all downstream analyses (Methods).

### Healthy volunteers and clinic patients show widespread differences in gene expression

To investigate overall gene expression patterns, we first tested for gene expression differences between all clinic patients (benign and cancer diagnoses) and healthy volunteers using nasal epithelium samples from both current and former smokers correcting for smoking status, pack-years, sex and age. We found extensive differences in gene expression between the healthy volunteer and clinic groups, with 5359 genes differentially expressed (FDR < .05, Methods). Genes showing increased expression in clinic patients were enriched for cilium assembly and organization, while genes showing reduced expression were enriched for oxidative phosphorylation and several immune-related pathways, such as neutrophil activation, antigen processing and presentation and response to interferon gamma (Sup. Tab. 3). When performing the same comparison in current smokers only, similar enrichment was found in the genes with increased and reduced expression. In former smokers who had quit for more than 1 year, there was no increased expression compared to healthy volunteers for genes related to ciliary function, but there was reduced expression of genes related to immune pathways such as inflammatory response, neutrophil activation and response to interferon gamma. These analyses demonstrate widespread expression differences betweenhealthy volunteers and clinic patients not solely aPributable to differences in smoke exposure, and suggest that an immunosuppressed state can be detected in the nasal epithelium of subjects from the clinic group during active smoking and for years after smoking cessation.

In contrast, comparing gene expression between patients with and without cancer in the clinic group and accounting for the same confounding (analysing current and former smokers together) yielded only 28 significantly altered genes (*Padj* < .05, Methods) in the bronchus, and no significantly differentially expressed genes in the nose. Among the 28 differentially expressed genes in the bronchus, 3 were up-regulated in patients with cancer: MMP13, a metalloproteinase known to increase lung cancer invasion and metastasis (*19*), EDA2R, a member of the tumour necrosis factor (TNF) receptor superfamily, members of which modulate immune response in the tumour microenvironment (*20*), and CTSL, a lysosomal cysteine protease involved in epithelial-mesenchymal transition (*21*). The 25 genes down-regulated in cancer patients were enriched in immune related GO terms, in particular neutrophil-mediated immunity (Sup. Tab. 4), consistent with our finding in the comparison between clinic patients and healthy volunteers in nasal tissue.

In summary, we observe major gene expression differences in nasal epithelium between healthy volunteers and clinic patients. However, we find no significant signal when comparing patients with cancer with those with a benign diagnosis within the clinic group. This result is in contrast to that obtained in the AEGIS study (*16*), which reported a notable difference in nasal gene expression between clinic-referred cancer and benign patients. However, we found a significant overlap between the set of differentially expressed genes between cancer and no-cancer in AEGIS and the set of differentially expressed genes between our clinic and healthy groups (*P* = 1.44×10^−5^). These results may be explained by subtle differences in the nature of the benign (non-cancer) diagnoses between the two studies. In our study, the majority of patients in the clinic group had clinical symptoms/imaging highly suspicious for lung cancer. Patients with a final benign diagnosis were predominantly due to significant typical bacterial infection/inflammation (pneumonia). However in the AEGIS cohorts many of the benign diagnoses, where known, were due to sarcoidosis, fibrosis, benign tumours or atypical infections (fungal and mycobacterial). Therefore, in our cohort the pre-test probablility for malignancy in the benign group was higher than in the AEGIS benign group.

### Gene expression response to smoke injury differs between healthy volunteers and clinic patients

Intrigued by these overall expression differences between volunteers and clinic patients we investigated the post-cessation dynamics of individual genes using a population-based approach. We first employed a Bayesian linear regression model to predict nasal gene expression in healthy volunteers as a function of smoking status, accounting for sex and age (Methods). This model classified genes as either *unaffected by smoking (US), rapidly reversible* (RR; no difference between former and never smokers), *slowly reversible* (SR; intermediate expression levels in former smokers compared to never and current), or *irreversible* (IR; no difference between former and current smokers). Additionally, genes were classified as *cessation-associated* (CA) if no difference was present between current and never smokers, but elevated or reduced expression was observed in former smokers (see Sup. Fig. 1 for a schematic).

In healthy volunteers 5755 genes were found to be affected by smoking status, out of which 513 genes show a strong effect (effect size > 0.4 for rapidly reversible, slowly reversible, irreversible genes, > 0.25 for cessation activated genes, Methods, Sup. Tab. 5). Most genes (485/513) were found to be rapidly reversible, in line with previous findings in bronchial tissue (*9*). GO pathway analysis of these genes revealed up-regulation of cellular detoxification, response to oxidative stress (e.g. CYP1A1, CYP1B1, AHRR, NQO1, GPX2, ALDH3A1) and keratinization (e.g. KRT6A, KRT13, KRT17, SPRR1A, SPRR1B, CSTA) pathways, and down-regulation of cilium organization (e.g FOXJ1, DNAH6, IFT81, CEP290, UBXN10), extracellular matrix organization (e.g. FN1, COL3A1, COL5A1, COL9A2) and interferon signaling (e.g IFI6, IFIT1, IFI44, RSAD2) in current compared to never smokers. Genes involved in inflammatory response were found both among the up-regulated (IL36A, IL36G, S100A8, S100A9, CLU) and down-regulated (SAA1, SAA2, IL33) genes. Principal components analysis using the rapidly reversible genes showed a clear separation of current smokers from all other subjects. In contrast, slowly reversible and irreversible genes placed patients on a trajectory from never smokers to current smokers, as expected (Sup. Fig. 2a).

We next repeated the above analysis on the clinic subgroup. In the absence of clinic never smokers, and since no technical or biological covariates could explain the observed overall expression differences between the groups (see Methods), we considered the healthy volunteer never smokers as a bona fide reference group for this analysis. We found 4112 genes with smoking-dependent expression changes, 584 of which showed a strong effect (same effect size thresholds as above, see Methods and Sup. Tab. 5). We evaluated this classification with a principal components analysis on the clinic subjects, similar to what was done for healthy volunteers, and found that patients clustered according to their smoking status, as expected (Sup. Fig. 2b). Of the 584 genes identified as dysregulated by smoke in the clinic patients, 233 were also found in the healthy volunteer analysis (*P* < .001, chi-squared test). However, while 227/233 were rapidly reversible in the healthy volunteers, only 112 of these 227 genes were also classified as rapidly reversible in the clinic group (Fig. 2a). Of the remaining 115 genes, one gene (BPIFA2) was now classified as irreversible and 22 genes as slowly reversible, including CYP1B1, a well-known detoxification gene, and BMP7, a gene previously shown to have a role in immunoregulation (*22*) (Fig. 2b). Ninety-two of 115 genes that were classified as rapidly reversible in the healthy volunteer group and as cessation-associated in the clinic group (e.g. UBXN10, Fig. 2b) showed a strong enrichment for cilia structure and function (Sup. Tab. 6). While cilia-associated genes were down-regulated in current smokers in both groups (consistent with cigarette smoke damaging airway cilia), the same genes showed increased expression in current and former smokers in the clinic group compared to the healthy volunteers. This observation in the clinic group might be linked to the decreased expression of interferon gamma-related genes in the clinic group, as it has been shown that interferon gamma suppresses ciliogenesis and ciliary movement (*26*).

**Figure 2:**
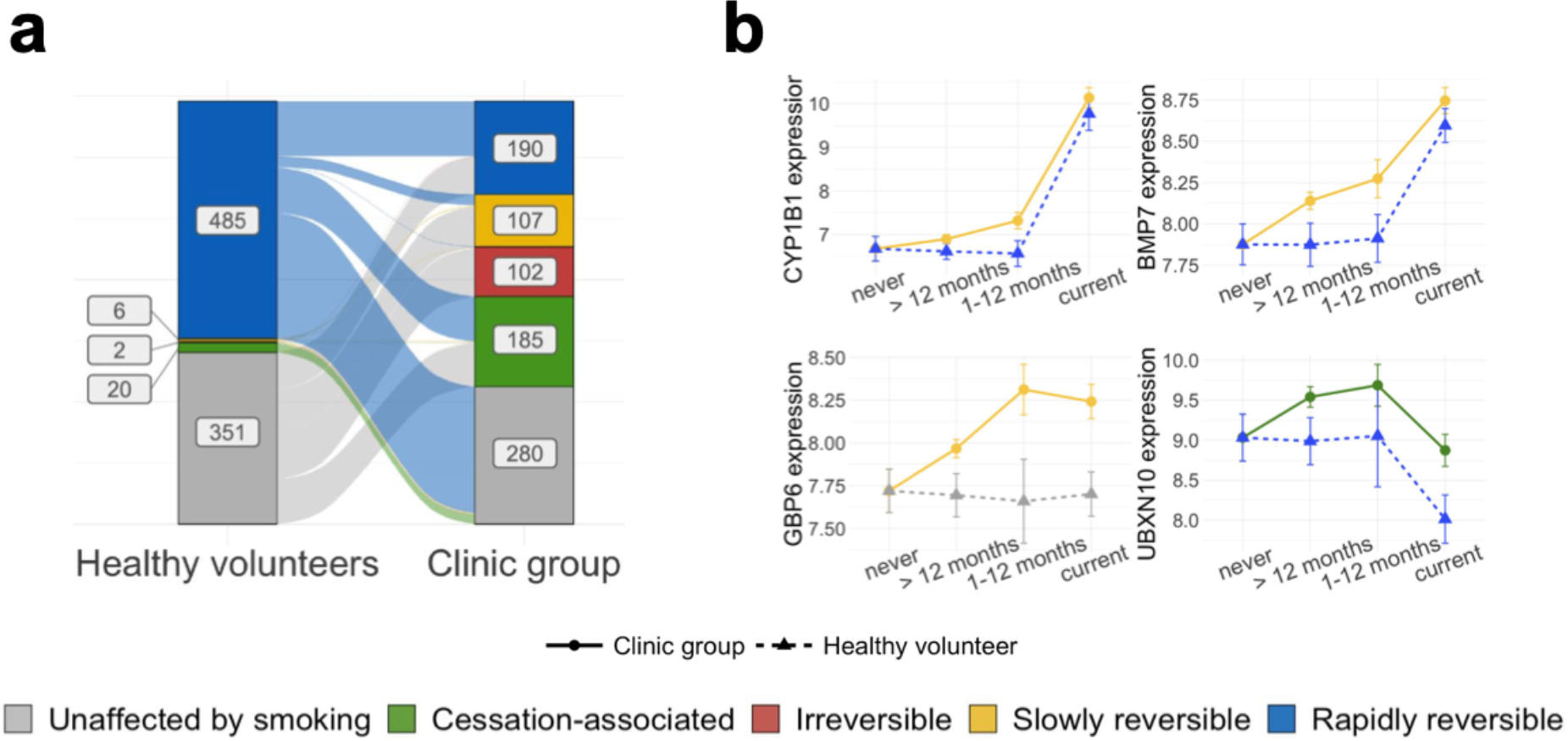
Smoke injury dynamics. **(a)** Plot showing a change of reversibility dynamics for the same genes in the healthy volunteer (left) and clinic (right) donor groups (genes classified as unaffected by smoking in both donor groups were removed); **(b)** Normalized gene expression over smoking status for 4 exemplar response genes with different post-cessation dynamics in the clinic and healthy groups, with linetype and shape representing donor status and colors representing the genes’ assigned reversibility classes.

Lastly, the 351 genes that showed smoking-dependent expression changes in the clinic group but not in the healthy volunteers (Fig. 2a) were strongly enriched in extracellular matrix organization and immune-related genes (including response to interferon gamma, neutrophil activation, chemotaxis and inflammation). For example, GBP6 showed down-regulation and slow reversibility in the clinic group (Fig. 2b) and is known to be associated with reduced overall survival in squamous cell carcinoma of the head and neck (*27*).

Overall, we observe striking differences in smoke-dependent gene expression in the clinic patients compared to volunteers that could not be explained by comorbidities or other covariates, with generally slower reversibility post-cessation in the clinic group. We hypothesise that some of the 749 genes with differences in smoke-dependent expression might reflect individual responses to the smoke injury and thus refer to them as *response genes*.

### Response gene expression levels predict disease status and may improve risk stratification for population screening

We postulated that the smoke-injury response genes we identified might provide evidence for a personalised smoke injury response and be candidate genes for a molecular biomarker of lung cancer risk. In the clinic group, where patients already show evidence of lung disease, such a biomarker would help identify patients with the highest need for further investigation. In the general smoker and former smoker population it could be added to existing methods of risk stratification to improve the identification of individuals who would most benefit from lung cancer screening thereby sparing those at lowest risk who would have least to benefit from screening.

Therefore, we trained two independent classifiers: a ‘clinic classifier’ that predicts the cancer status of each sample (cancer vs clinic benign and healthy volunteers: potentially of use in the clinic), and a ‘population classifier’ that predicts the donor group that the samples were taken from (clinic benign or clinic cancer vs healthy volunteers: potentially of use in risk stratification for population screening). For both classifiers, we used gene expression data from the 749 response genes together with clinical information (sex, age, smoking status and pack-years; see Methods) in a lasso-penalized multivariate logistic regression, and derived a log-odds score from each classifier. In line with the observed strong expression differences between healthy volunteers and clinic patients, the ‘population’ score clearly separates healthy volunteers from clinic subjects (Fig. 3a). Interestingly, the ‘clinic’ score (Fig. 3b) additionally distinguishes the benign and cancer patients within the clinic group, placing benign subjects between healthy volunteers and cancer subjects. As expected, the two scores are highly correlated (Pearson correlation = 0.8, *P* < .001, Sup. Fig. 3a). Both scores yielded high area under the curve (AUC) values for both precision-recall (clinic score: mean AUC-PR=0.83 after 10-fold cross validation; population score: mean AUC-PR=0.85, 10-fold cross validation, Fig. 3c-d) and receiver-op-erator characteristics (clinic score: mean AUC-ROC=0.84, 10-fold CV; population score: mean AUCROC=0.92, 10 fold CV, see also Methods) and performed significantly better than a model using the same number of randomly selected genes (Sup. Fig. 4). In practice, to reach a sensitivity of 95% for the population score, one would use a score threshold of 2.69, that would result in an average false positive rate of 42.8%, while to reach a similar sensitivity using clinical data alone would result in a false positive rate of 74.5%. For the clinic score, a score threshold of -1.46 gives a 95% sensitivity and false positive rate of 62.1%, while similar sensitivity with clinical data alone would result in a false positive rate of 67.8% (Fig. 3c-d). These results indicate that models incorporating gene expression data of the response genes defined above performed significantly better than models built on clinical covariates alone (see also inset of Fig. 3c-d for a comparison of the performance of models based on gene expression data alone, clinical covariates alone or a combination of gene expression data and clinical covariates). In addition, both scores retained their ability to separate the patient groups after regressing out all potential confounders, confirming that gene expression data improves classification compared to using clinical covariates alone (Sup .Fig. 3b-c).

**Figure 3:**
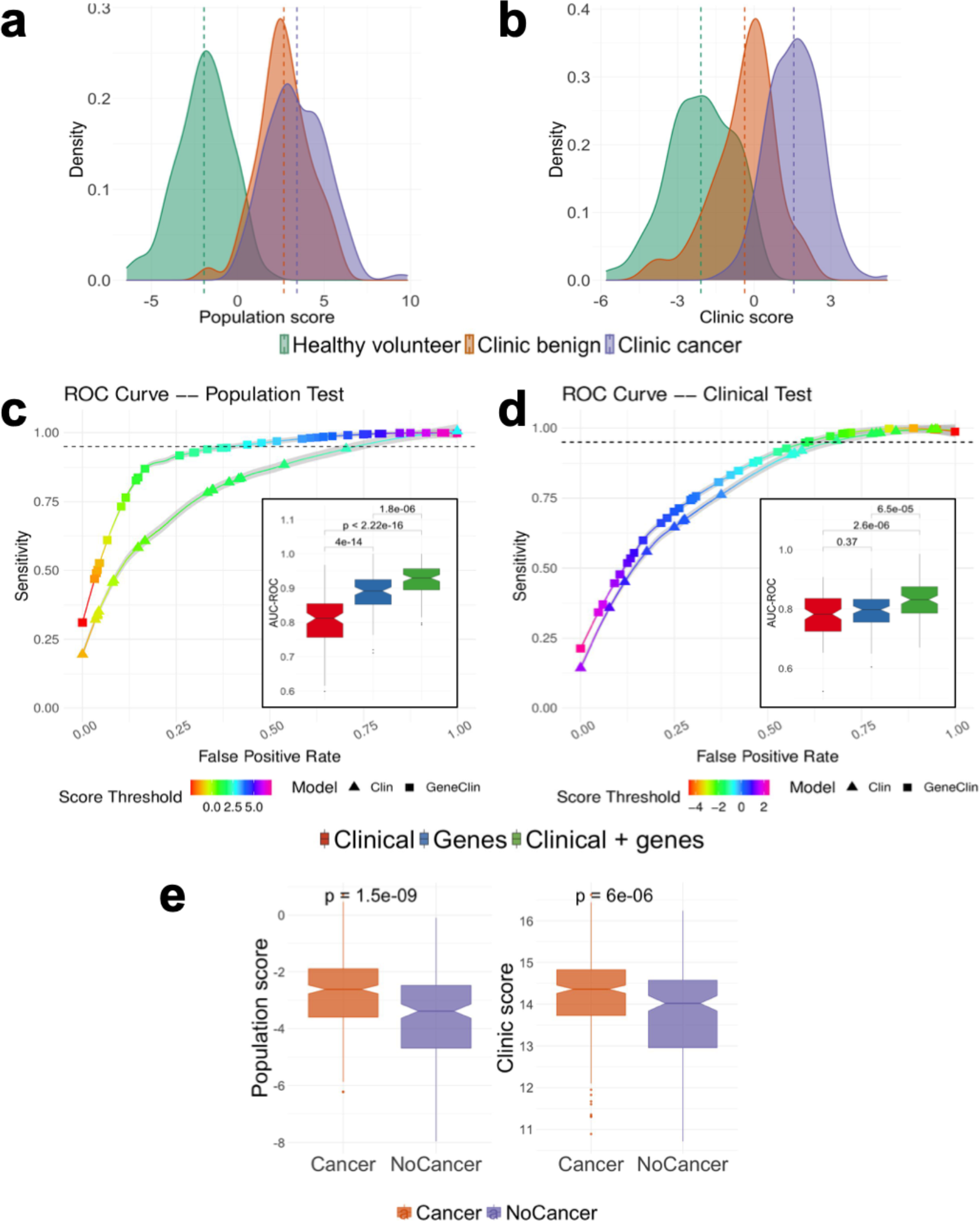
Disease status prediction based on response genes. **(a & b)** Risk score distribution for the population test **(a)** and the clinic test **(b)** predicted from the clinical variables and the expression of the response genes using a penalized regression (See Methods). The risk distributions are presented separately for healthy volunteers (green), clinic patients without cancer (orange) and clinic patients with cancer (purple) **(c & d)** ROC curves for the population **(c)** and clinic (**d**) scores. For each case, we present the ROC curve for the model trained on clinical data (triangles) or on gene expression and clinical data (squares). Each curve is an average obtained across 100 cross validation (CV) experiments and the grey area surrounding the curve gives the standard error. The color of the curve represents the test threshold corresponding to the represented Sensitivity / False Positive Rate compromise. (**Inset)** Area under the ROC curve, in 100 CV rounds, for a clinical-only model (red) the model constructed on the response genes (blue) and a model constructed on a combination of clinical information and response genes (green) for the population **(c)** and clinic **(d)** classifiers. **(e)** The population and clinic classifiers applied to nasal samples from the AEGIS cohort.

We also assessed the performance of the trained population and clinic risk score models separately on current and former smokers. We found that the population risk score is equally applicable to current and former smokers: a significant difference in the risk score of the healthy volunteers and clinic subjects can be observed, even after regressing out clinical covariates and confounding (Sup. Fig. 5). While the clinic risk score performs well on both groups, the added value from gene expression data appears less important in the clinic score, in particular in former smokers (Sup. Fig. 5). Finally, we find that our classifiers are efficient at separating subjects regardless of their cancer stage, cancer type (squamous carcinoma or adenocarcinoma), and COPD status (Sup. Fig. 6), and that our classifiers capture differences in risk that persist for more than 10 years after smoking cessation (Sup. Fig. 7).

Finally, we validated our classifiers by applying them to an independent cohort. No publicly available cohort matches the composition of our cohort, in particular because of the absence of a healthy group of current and former smokers distinct from the clinic-referred patient group. However, the AEGIS cohort (*28*) includes nasal samples from clinic-referred patients with pulmonary nodules and a diagnosis of lung cancer or benign disease. We applied our two classifiers to this cohort, and found a good separation between subjects with and without cancer, despite the different gene expression quantification technologies and populations of origin of the patients (Fig. 3e, Sup. Fig. 8). We found a stronger separation between patients with and without cancer using the AEGIS nasal classifier from Perez-Rogers et al (2017) (*16*) on the AEGIS data (Sup. Fig. 8a). However, we note that the AEGIS classifier (*16*), when applied to our data, mostly differentiates healthy volunteers and clinic patients while the difference between the scores of the cancer and no-cancer patients is only modest (Sup. Fig. 8b). These results confirm the ability of our classifier to stratify patients, even when applied to patients from different clinical contexts.

Overall, our results demonstrate that classifiers based on nasal gene expression have the potential to improve risk stratification of current and ex-smokers in both a population screening context and a clinic context.

### Alterations in immune pathways underlie the lung cancer risk classification

To gain insights into the mechanisms of risk, we asked which genes robustly contributed most to the classifiers by identifying genes selected in more than 80% of the cross validation (CV) rounds (Sup Fig. 9). Among the 46 genes selected most often in either of the risk prediction models, we found genes that were previously identified as important players in lung cancer development, e.g. SAA2 [18], HAS2 (*29*), (*30, 31*) or TGM3 (*32*–*35*), in line with the current literature.

However, the genes used as predictors of risk in our model reflect a wide variety of smoking-associated alterations. In order to gain some mechanistic insight, we investigated risk contribution at the pathway level. First, we performed GO enrichment analysis on the list of smoke injury genes (both the ones identified in the healthy volunteers and in the clinic group) to identify the main pathways affected by smoke. We found that the smoke injury genes are mainly involved in xenobiotic metabolism and response to oxidative stress, extracellular matrix organization, keratinization, ciliary structure and mobility, and immune response (Sup. Tab. 7). We then chose 8 GO terms as represen-tatives of these alterations: *Keratinization, Extracellular matrix organization, XenobioIc metabolism, Cilium organization, Inflammatory response, Neutrophil mediated immunity, Response to interferon gamm*a, and *Antigen processing and presentation*. We calculated geneset metascores for each of these GO terms (Fig. 4a and Sup. Fig. 10). For some of these pathways, such as Keratinization, we observed a similar, rapidly reversible dynamic in healthy volunteers and clinic patients (Sup. Fig. 10a). For most pathways, however, the dynamics were different in the two donor groups. *Cilium organization* appeared to be rapidly reversible in healthy volunteers, while in clinic patients it showed increased expression in former smokers, with no difference between current and never smokers. *XenobioIc metabolism* showed a slower reversibility in clinic patients than healthy volunteers (Sup. Fig. 10a). For all immune-related pathways, we observed reduced expression in current smokers, and a slow reversibility dynamic, uniquely in clinic patients (Fig. 4a); we also observed that their activity does not revert to healthy never-smoker level even long after smoking cessation (Sup. Fig. 10b).

**Figure 4:**
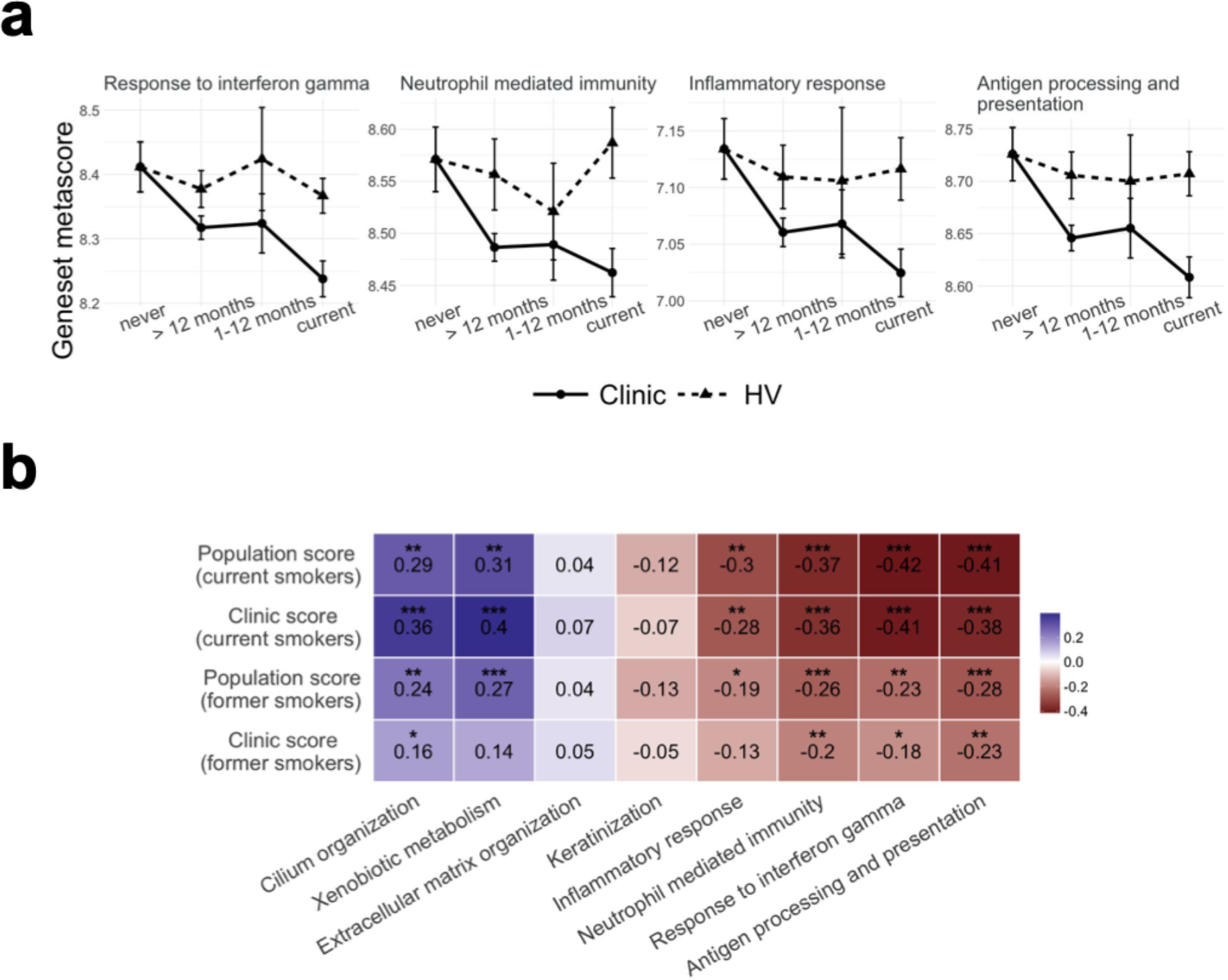
Pathway analysis and contribution to risk. **(a)** Comparison of geneset metascore over smoking status for 4 immune-related GO terms in healthy and clinic subjects. **(b)** Correlation between the population or clinic risk score and geneset metascore for the 8 gene sets representing biological functions altered by smoking; correlation is shown separately for current and former smokers (> 12 months); Spearman correlation values are reported, as well as the associated p-values (*: *P* <= 0.05, **: *P* <= 0.01, ***: *P* <= 0.001).

To identify which of these pathways contributed most to increased risk, we then calculated the correlation between geneset metascore in each subject and subjects’ risk scores from the population and clinic classifiers. We calculated these correlations for current and former smokers (> 12 months) separately, to be able to identify differences in geneset contribution to risk in the two groups that might reflect differences between acute smoke injury response and the long-term consequences of past smoke exposure (Fig. 4b). In current smokers, while *Keratinization* and *Extracellular matrix organization* did not significantly correlate with either risk score, the remaining four genesets tested showed moderate but significant correlation with both risk scores, pointing to alterations of the xenobiotic detoxification pathways, ciliary function and immune response as major contributors to patient-specific differences in risk. In former smokers, the population risk score correlated with the same 4 GO terms indicating that detoxification pathways, ciliary function and immune response are the main contributors to overall risk of lung disease. In contrast, only pathways related to immune alterations (*Response to interferon gamma, Neutrophil-mediated immunit*y, *Antigen processing and presentation*) correlated with the clinic risk score in former smokers, while no correlation was observed with *XenobioIc metabolism*, and only a very weak correlation with *Cilium organization* (Fig. 4b). These results indicate that immune alterations are significant contributors to the risk of cancer in both current and former smokers in the clinic group.

### Patient-specific genetic background modulates the smoke injury response

Germline genetic variation may influence individual differences in response to airway smoke injury, and hence, risk of smoking-related lung cancer. To investigate this, we first conducted an eQTL analysis on nasal and bronchial epithelium separately and jointly to identify variants that affect the expression of neighbouring genes (Methods). We obtained 990 (bronchial), 1316 (nasal) and 1695 (combined) eQTL effect genes (e-genes) at 1% FDR. We found a significant overlap between the nasal and bronchial e-genes (Sup. Fig. 11a), with 574 genes in common (corresponding to 58% and 44% of the bronchial and nasal eQTL respectively, Fisher’s exact test *P* < .001). Similarly, we found a correlation of 0.56 between the adjusted p-values of the lead variants between both sets (Sup. Fig. 11b), confirming shared *cis*-regulation between the nasal and bronchial epithelium.

To further study the interaction between subject-specific genetic background and environmental factors we next leveraged this eQTL catalogue to search for genetic variants within the 749 response genes that might modulate gene expression differently depending on subjects’ smoke exposure. We identified 78/749 genes with at least one lead eQTL variant with genome wide significance at 10% FDR, (Sup. Tab. 8). We then tested for an interaction effect between smoking status and genotype for all 78 lead eQTL variants on gene expression. We identified 11 genes (CH25H, LHX6, WNT5A, DRAM1, SULF1, LGALS7B, HAPLN4, FXYD5, EFCAB2, TOX and SPRR1A, see Sup. Fig. 12) whose expression changes in response to smoke are modulated by the presence of genetic variants (nominal *P* < .1, Sup. Tab. 8), suggesting that those genetic variants might modulate the response to smoke injury and to lung cancer risk. For example, up regulation of FXYD5 has been shown to correlate with tumor size (*36*) and poor survival (*37*) in NSCLC and to be implicated in many cancer types as FXYD5 enhances NFκ-B transcriptional activity, promotes angiogenesis and increases tumor cell’s migration and invasion abilities (*38*). Finally, this protein also promotes inflammation in epithelial cells, notably in lung tissues (*39*). Analysing the expression of this gene in our cohort, we find that subjects with a homozygous reference genotype at the *19:35660670:G:A* locus have similar levels of expression both in never, ex, and current smokers (Fig. 5a). On the contrary, subjects that have a heterozygous or homozygous alternative genotype present higher levels of expression of this gene in response to smoke (Fig. 5a), which might increase their lung cancer risk. We observe similar trends for the 10 other response genes stated above (Sup. Fig. 12, Sup. Tab. 8). This finding demonstrates how subjects’ specific genetic background can influence their reaction to cigarette smoke and in turn might affect their risk of developing lung cancer.

**Figure 5:**
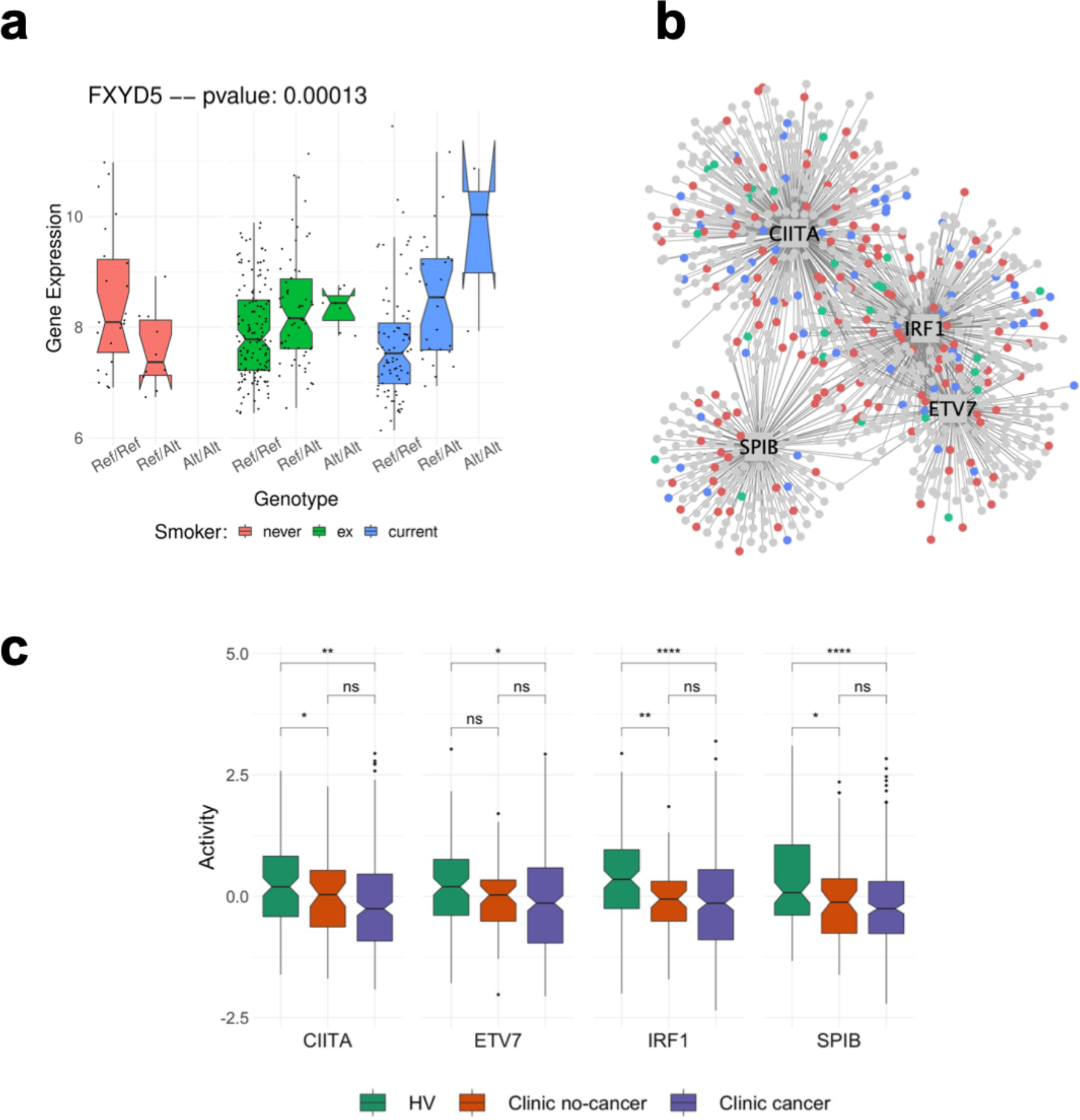
Genotype background influences lung cancer risk. **(a)** Combined environmental and genetic effect on the expression of the FXYD5 gene in nasal tissues. For each nasal sample, we present the expression level of the gene FXYD5 separately for never (pink), former (green) and current (blue) smokers. Samples are further stratified depending on the genotype of the subject at the 19:35660670:G:A locus (Ref/Ref: homozygous reference; Ref/Alt: heterozygous; Alt/Alt homozygous Alternative). The p-value gives the significance level of an interaction effect of the smoking status and the genotype at the 19:35660670:G:A on the expression of the FXYD5 gene (see Methods). **GWAS enrichment analysis: (b)** Network representation of the 4 bronchial regulons enriched in GWAS genes. The 4 TFs are shown as squares and their targets in the bronchial network as circles. Color of the nodes indicates whether the gene/TF is a smoke injury risk gene **(blue)**, a gene that co-localizes with a GWAS hit (i.e. no threshold on eQTL significance) **(red)** or both **(green)**. The level of overrepresentation for genes in the network of those TFs can be found in Table 2 (for the GWAS) and Table 3 (for the response genes); **(c)** Activity level of each of the 4 TFs in nasal tissue, depending on the disease status of the patient.

### Common germline variants regulate interferon gamma genes and are linked to known lung cancer risk loci

We next identified GWAS hits that were in strong linkage disequilibrium in the UK population to SNPs that we found to be regulating the expression of nearby genes in our eQTL analyses (Methods). Among the 1261 GWAS lung cancer risk loci, our analysis identified 63 GWAS risk loci from 13 different studies with variants that significantly affect the expression of a nearby gene at a 5% FWER threshold (Sup. Tab. 10). These 63 eQTL/GWAS variants were linked to the expression of 41 genes, notably including 10 genes implicated in the interferon gamma signalling pathway. Pathway enrichment confirmed a strong enrichment for genes involved in response to interferon gamma (hypergeometric test, P*adj* = 7 × 10^−13^), as well as for other immune-related functions (e.g. *innate immune response, anIgen processing and presentation of exogenous pepIde anIgen, regulation of immune response, T cell receptor signalling pathway*; see Sup. Tab. 11 for the full list of enriched GO terms).

To better understand the mechanisms by which GWAS variants might increase lung cancer risk, we looked for a link between 41 genes linked to a GWAS risk locus and transcriptional regulatory network in bronchial tissue. To do so, we inferred a TF-target interaction network from bronchial expression data (see Methods) and searched for TFs whose targets were enriched for the 41 genes. We found 4 TFs showing a strong enrichment (hypergeometric test, *Padj* < .05, see Methods), ETV7, SPIB, IRF1 and CIITA (Fig. 5b), all of which are known players in the interferon gamma mediated signalling pathway (*40*–*43*). We further confirmed the enrichment of GWAS genes in these 4 TFs by using a wider list of GWAS genes with a relaxed eQTL cut-off (nominal *P* < .05), and still found a 2- to 3-fold enrichment in all 4 TFs (Table 1). Analyzing the activity of those 4 TFs in the nasal samples, we found significant differences between healthy volunteers and clinic patients, in particular a lower activity in clinic patients, confirming the importance of these 4 TFs in the progression toward a disease status (Fig. 5c, and see Sup. Fig. 13 for the activity of the same 4 TFs in the bronchial samples of clinic patients with and without cancer). In contrast, we found that the levels of activity of those 4 TFs were similar in clinic patients with and without cancer (Fig. 5c and Sup. Fig. 13). We further tested whether our set of response genes was enriched within the targets of those TFs and indeed found that all 4 TFs are enriched for response genes (2 to 3-fold enrichment, nominal *P* < .05, Table 2).

**Table 1:** Overrepresentation of GWAS associated genes in the regulatory network of four TFs. Regulon size: the number of genes in the regulatory network for each TF; **# hit:** number of genes, among each TFs regulatory network that we annotate as a GWAS-linked gene (in parenthesis: expected number of GWAS genes in the regulatory network of the TF); **FDR:** False discovery rate of the overrepresentation of GWAS hits in the TF regulatory network (hypergeometric test, see methods). Each test is performed for 3 sets of genes defined using a hard (*P* <1e-06; 44 genes); lenient (*P* < .05; 569 genes) or no threshold (3181 genes) on eQTL significance levels.

| GWAS Genes |  |  |  |  |  |  |  |
| --- | --- | --- | --- | --- | --- | --- | --- |
|  |  | Hard Threshold |  | Lenient Threshold |  | No Threshold |  |
| TF | Regulon Size | # Hit | FDR | # Hit | FDR | # Hit | FDR |
| **IRF1** | 318 | **10**<br>(1.5) | 2.9 1e-07 | **34** (12.5) | 2.0 1e-07 | **83** (60) | 0.07 |
| **CIITA** | 372 | **9** (1.5) | 2.4e-08 | **26** (13) | 0.08 | **83** (69) | 1 |
| **SPIB** | 174 | **6** (0.5) | 2.9e-04 | **14** (5.8) | 0.6 | **43** (30.7) | 1 |
| **ETV7** | 171 | **4** (0.8) | 0.088 | **14** (5.4) | 0.5 | **35** (28.7) | 1 |

**Table 2:** Overrepresentation of the response genes in the regulatory network of four TFs. Regulon size: the number of genes in the regulatory network for each TF; **# hit:** number of genes, among each TFs regulatory network that we annotate as a risk gene (in parenthesis: expected number of GWAS genes in the regulatory network of the TF); **P-value:** the p-value of the overrepresentation of response genes in the TF regulatory network (hypergeometric test, see methods).

| Response Genes |  |  |  |  |
| --- | --- | --- | --- | --- |
| TF | Regulon Size | # Hit | P-value | FDR |
| **IRF1** | 318 | **37** (11.5) | 5.19 1e-11 | 7.67 1e-08 |
| **CIITA** | 372 | **45** (13.2) | 1.31 1e-13 | 1.96 1e-10 |
| **SPIB** | 174 | **14** (6.16) | 0.0013 | NS |
| **ETV7** | 171 | **14** (6.08) | 0.0011 | NS |

Altogether, these findings suggest that the effects of inherited variation on lung cancer risk may be exerted in part through a different immune response following smoke injury, creating an immunosuppressed environment that favours the final steps to the emergence of a cancer.

## Discussion

In this study, we demonstrate that gene expression data from nasal epithelium has the potential to improve lung cancer risk stratification within the general population of current and former smokers. Using healthy never smokers as a baseline, we have compared smoking-dependent patterns of gene expression between healthy volunteers and clinic patients under investigation for lung cancer. We have developed gene-expression-based classifiers to separate these groups, thereby revealing striking differences in the long-term persistence of gene expression patterns after smoking cessation. Using pathway analysis, we have inferred the mechanisms that underlie these differences. We found that known lung cancer risk loci regulate the expression of genes that are enriched in specific pathways that were also deregulated in response to smoking. These pathways include neutrophil mediated immunity and response to interferon gamma, suggesting that immune dysregulation is causally involved in the aetiology of non-small cell lung cancer. Our results are consistent with recent studies linking immune-related genetic variants to a variety of lung-related phenotypes (*44*). Together, they support and extend the model in which genetically influenced differences in immune regulation interact with smoking and other injuries, including air pollution (*45*), to create an airway cellular environment which is associated with impaired lung function and an increased risk of lung cancer. Our population classifier is, to our knowledge, the first gene-based classifier to address specifically risk stratification for lung cancer in the healthy smoker population. With an average cross-validated AUC (ROC) of 0.92 (Fig. 3a,c), the classifier identifies 95% of high risk individuals with a false positive rate of around 40%. The gene expression data adds to the power of the classifier over clinical data alone (Fig. 3c). If confirmed, these results suggest potential value from including gene expression data in such a classifier as population-based lung cancer screening becomes more widely adopted.

It is important that the classifier be validated in the precise clinical context in which it will be applied. However, no suitable data set for validation is currently available. We suggest that our results are sufficient to support inclusion of a confirmatory study, possibly including both rederivation and validation, in the lung cancer screening initiatives currently under development. In such studies, the contribution of gene expression data should be assessed alongside other potential predictors.

Support for the validity of our classifiers and thus for these further studies comes from two sources. First, our cross-comparisons with the AEGIS dataset (16) which showed that our classifiers have power to discriminate patients with and without cancer within that dataset (Fig. 3e), even though the cohort was microarray-based, and the samples derived from a different clinical context. Second, we show (Fig. 4) that the genes that contribute most to the classifiers belong to pathways related in particular to inflammatory and immune function, which are in turn linked to genetic variation at lung cancer GWAS loci. This is evidence for a causal role of these genes in lung cancer risk. Consistent with this, our classifiers are equally efficient at identifying individuals with early or late stage disease (Sup. Fig. 5c), and in predicting squamous or adenocarcinoma (Sup. Fig. 5b). The classifiers are effective in both current and former smokers, and the clinic patients (cancer and benign) continue to show an elevated risk score ten years and more after stopping smoking (Sup. Fig. 8), consistent with the known persisting cancer risk in former smokers. This may allow identification of those former smokers most at risk, and in time, open up approaches to lowering that risk based on the mechanisms involved in that individual.

Using a geneset metascore analysis, we identified immune-related pathways, in particular response to interferon gamma and antigen processing and presentation, as the pathways that contribute most to our lung cancer risk scores (Fig. 4b) in both current and former smokers. IFN-***γ*** is a molecule that is involved in anti-tumor immune response by activating cellular immunity and exhibiting anti-proliferative, pro-apoptotic and anti-angiogenic properties within the tumour microenvironment (*46*). An immunosuppressive state favoured by the decreased expression of genes involved in IFN-***γ*** signalling and antigen presentation was observed both in lung cancer and in bronchial premalignant lesions, and suggested to promote the progression to invasive disease (*18, 47*). We observe these alterations in healthy-appearing nasal tissue affected by the smoking-associated field of injury, suggesting that this immunosuppressive, cancer-promoting, state is present at even earlier steps of carcinogenesis.

Our analysis based on the known NSCLC GWAS risk loci provides the critical causal links between risk variants and the activity of four transcription factors known to be active in interferon gamma signalling (CIITA, ETV7, IRF1, SPIB). We also identified 10 genes whose response to smoke differed between healthy and clinic subjects and whose expression was regulated by a gene-by-environment interaction between the genetic background of subjects and their smoking behaviour (Fig. 5a, Sup. Fig. 12). While these results demonstrate how genetic background can affect individual response to smoke injury, and so lung cancer risk, larger cohorts will be needed to explore systematically the interaction between smoking behaviour and individual genetic background genome-wide. This may in time uncover differences in mechanisms of risk between individuals, and allow risk-lowering interventions to be tailored appropriately.

Together, our results suggest a model for smoking-related lung cancer risk in which genetically determined differences in the immune and inflammatory responses to cigarette smoke and other environmental exposures modulate the bronchial cellular environment and increase the probability of progression towards cancer. The persisting risk in former smokers is, at least in part, driven by the persistence of this altered cellular environment. Individuals exhibiting altered cellular environments are both at higher risk and more likely to be symptomatic and to attend the respiratory clinic than others, whether they have cancer or not; hence the incomplete separation between benign and cancer within the clinic group.

Recent papers (*17, 18, 48*) have investigated the role of altered immune responses in the progression of preneoplastic airway lesions. Their findings will lead to better prediction and intervention in the management of patients already deemed at sufficient risk to justify bronchoscopic surveillance. Our study adds to that of Kachuri et al (*44*) by extending knowledge of the mechanisms that link risk to impaired lung function at the earlier stages of smoking-related lung cancer development, with implications for risk prediction, screening, and eventually strategies for risk reduction.

## Methods

### Availability of the code and data

Scripts that were used to conduct the analysis presented in this paper are available in the bitbucket repository accessible with the following link:

https://bitbucket.org/schwarzlab/paper-debiase-massip-2021

Raw and processed data will be available upon acceptance of the paper on the EGA website.

### Cohort and sample collection

487 donors were recruited into the CRUKPAP cohort at Royal Papworth Hospital, Cambridge (UK), including 114 healthy volunteers (HV) and 337 patients being investigated for suspicion of lung can-cer. The eligibility criteria for healthy volunteers were: age 18 or above; current or former smokers must have smoked at least 100 cigarettes in their lifetime; exclusion of individuals with previous history or current suspicion of airway or lung cancer; exclusion of individuals with ‘bleeding disorders’.

All participants were stratified into smoking cessation categories as follows: 45 never smokers (NV), 289 former smokers (FS) and 153 current smokers (CS). Former smokers were further divided into categories: > 1 year after cessation (FS1, n=234), 1-12 months after cessation (FS2, n=45) and < 1 month after cessation (FS3, n=10). Smoking status for all subjects was confirmed via blood cotinine test. Cumulative smoke exposure measured in pack years was recorded and stratified into four categories: ‘none’ (PY1), < 10 years (PY2), 10-30 years (PY3) and > 30 years (PY4). For suspected lung cancer patients, both COPD status and final cancer diagnosis (lung cancer / no lung cancer) were recorded.

From these donors 413 nasal epithelial curettages were collected using Arlington Scientific ASI Rhinopro nasal curettes. Briefly, the nostril is opened with a nasal speculum to identify the inferior turbinate. Under direct vision the tip of the nasal curette is gently scraped over the turbinate to obtain a ‘peel or curl’ of epithelial tissue. The curl of tissue is then removed by flicking the curette while the tip is submerged in RNAlater™ collection medium and presence of the curl floating in the medium is confirmed by visual inspection. This procedure is repeated twice for each nostril per donor. RNA integrity (RIN) was checked for all samples and we found >80 % of samples to have RIN 6 or better.

Bronchial brushings were collected using 2.0mm brush diameter cytology brushes (Olympus Medical, UK) from 236 patients undergoing flexible bronchoscopy as part of investigations for suspected lung cancer.

For 162 donors, both nasal and bronchial samples were available. Sample collection and diagnosis took place contemporaneously. All samples underwent short-read RNA sequencing using Illumina TruSeq library generation for the Illumina HiSeq 2500 pla{orm. Blood samples were taken from 467 donors and germline genotyped using the Illumina Infinium Oncoarray pla{orm at 450K tagging germline variants. Total gene expression levels (TPM and variance stabilised) were determined for 18,072 protein coding genes for all samples using DeSeq2.

Research ethics approvals for sample collection from participants in this study were given by East of England Cambridge Central REC 13/EE/0012 and the National Research Ethics Service Committee South East Coast – Surrey 13/LO/0889.

### RNA extraction and sequencing

Tissue samples from bronchial brushings and nasal curettes were stored in 500μl RNALater overnight at 4oC, and then at -80oC for longer-term storage. RNA was extracted using Qiagen MiRNeasy columns according to the manufacturer’s protocols. Briefly, bronchial brushes were rinsed in PBS, brushes transferred into 700μl Qiazol and cells lysed by vortexing twice for 30 seconds. For nasal samples the RNALater containing nasal tissue (500μl) was diluted with 2ml of PBS and spun at 10,000 rpm for 10 min. The cell pellet was lysed by resuspension in 700μl Qiazol. For both types of samples, the Qiazol lysate was applied to a QiaShredder tube (#217004) and spun at 13,000 rpm for 2 mins. The homogenate was kept at room temperature for 5 mins, followed by chloroform extraction using PhaseLock tubes. Nucleic acids in the aqueous phase were precipitated using 1.5 volumes of 100% ethanol and DNA was digested using DNAse I. Finally, RNA was isolated from the mixture using RNAeasy mini spin columns. RNA was quantified using a Qbit measurement and quality assessed us-ing an Agilent Bioanalyzer. For samples with a RIN greater than 7, a total of 500ng of RNA was used for Illumina TruSeq Library generation. Sequencing was carried out on HiSeq 2500 Illumina sequencers. Sequencing was carried out in two separate multiplexed experiments.

### RNA sequencing data processing

Quality control using FastQC showed good sequence quality and no adapter contamination for all samples. Alignment was carried out with TopHat2, using as reference the human genome version GRCh37. Read counts were computed for all protein-coding genes with *subread featureCounts v1*.*6*.*0*. The data was produced in 2 experimental batches, producing a strong batch effect that can be observed in the raw data. Moreover, a group of samples from one of the batches has lower total counts compared to the other samples (Sup. Fig 14a).

Raw counts were normalized using *DESeq2*’s variance-stabilizing transformation, which had the advantage of partly correcting the previously mentioned batch effects (Sup. Fig 14a). Genes with across-samples log variance smaller than -4 were discarded from further analysis. Total gene expression levels (variance stabilised) were determined for 18,072 protein coding genes for all samples. To ascertain that experimental batch did not covary with any clinical covariate, we computed the strength and significance of association between batch and the other covariates using Cramer’s V and chi-square test. We did not observe a significant association between batch and age, sex, COPD, smoking status, pack-years and donor population of origin (healthy volunteer/clinic patient). We only observed a weak but significant association with cancer status (Sup. Fig 14b).

To assess the overall contribution of clinical and environmental variables to gene expression in the nasal epithelium, we also extracted variance components using a linear model, regressing donor population of origin (healthy volunteer/clinic patient), cancer status, smoking status, pack-years, sex, age, COPD and experimental batch against total gene expression across all genes (Sup. Fig 14c). We found that donor population of origin and smoking status contribute most to gene expression variability (28.8 and 25.4% of total explained variance. Notably, donor population of origin still contributes significantly to the explained variance after accounting for all other clinical and technical covariates.

### Differen3al expression analysis

All differential expression analyses were performed with *DESeq2 v1*.*26*.*0*. Age, experimental batch, sex and pack-years were included as confounding variables. Genes with multiple-testing-adjusted (Benjamini-Hochberg) p-values < 0.05 were considered differentially expressed. For differential expression between clinic cancer and clinic benign in bronchial samples, 8 genes had artificially high (>20) absolute fold-change, due to their very low average expression across samples. These genes were removed from the list of differentially expressed genes.

### Modelling time-dependent dynamics of smoke injury in nasal 3ssue

#### Gene expression dynamics

To identify genes affected by smoke and characterize their post-cessation expression dynamics, we applied Bayesian linear regression and model selection (R package *BAS v1*.*5*.*3*). We modeled the expression of each gene on smoking status, where smoking status is encoded in 3 variables:

- CS (0/1) indicating current-smoker status
- FSS (0/1) indicating former-smoker status
- FS (0/1/2/3) indicating time since smoking cessation

Additionally, the model includes age, sex and experimental batch as confounding variables.

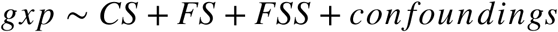

We tested for inclusion of each of the variables into our model and inferred posterior probabilities for all eight possible models to retrieve the most likely time dynamic of gene expression changes for each gene individually. Each combination, or group of combinations, corresponds to a gene class among *unaffected by smoking, rapidly reversible, slowly reversible, irreversible* and *cessation-associated* (Sup.Fig.1). Each gene is assigned to the class with the highest posterior probability. To identify genes for which smoking has the strongest effect, we applied a threshold on the beta coefficient, and retained only genes with a beta CS greater than 0.4 for rapidly reversible, slowly reversible and irreversible genes, and beta FSS greater than 0.25 for cessation-associated genes.

### Derivation of population and clinic risk scores

L1-penalized multivariate logit regression was performed with R package *glmnet 3*.*0-2* using only the nasal gene expression data. Patients status was encoded with a binary variable (cancer: 1; no cancer 0 for the clinic classifier; Clinic patient: 1, Healthy Volunteer: 0 for the population classifier), and patients with *Ineligible* status were excluded from the analysis. In the gene expression classifiers, the status of each patient was predicted based on the expression of the 749 response genes and 4 clinical covariates, namely sex, age, smoking status and packyears, all of which were encoded as numerical variables (smoking status encoding: Never smokers: 0, Ex >1year: 1; Ex 1-12months: 2; Ex <1m: 3, current smokers: 4). For the clinical classifier we also used a lasso regression, using only sex, age, smoking status and packyears as predictors. The lasso shrinkage parameter (λ) was chosen to minimize the mean cross-validated error (“lambda-min” option in the cv.glmnet function). Area under the receiver operating characteristic curve and precision recall curves were computed using the PRROC package, after 10 rounds of 10-fold cross validation experiments. To compare performances of the response genes to performances on random genes, we randomly drew 20 sets of 749 genes among the 18,072 protein coding genes retained for all analyses, and cross validations experiments were conducted on the same test and training set as the one used with the response genes.

### Gene ontology analysis and pathway analysis

All Gene Ontology (GO) enrichment analyses were performed using *clusterProfiler v3*.*14*.*3*. GO terms with adjusted (Benjamini-Hochberg) p-values < 0.05 were considered enriched.

Pathway metascores were calculated by averaging vst-normalized gene expression of genes belonging to the selected genesets, after regressing out experimental batch effect.

### Genotyping data pre-processing

#### SNP phasing and imputation

We phased the 450,000 germline genotypes using a statistical phasing algorithm (*eagle* v2.4.1) and population data from the 1000 genome project. For each haplotype, we then imputed missing genotypes using the *minimac4* pipeline. This allowed us to impute the genotype of each subject at 46,000,000 positions. After filtering out SNPs with low imputation quality (Rsq<0.8), we were left with 7,650,214 SNPs in total for each sample.

#### LD Pruning

First, we only considered SNPs that have a minor allele frequency greater than 1% in our cohort, reducing the number of SNPs to 5,772,170. Next, we removed SNPs in strong LD. To do so, we filtered out SNPs with a Variant inflation frequency larger than 20, with VIF= 1/(1-r^2). This threshold thus corresponds to removing SNPs with a multiple correlation >0.95. VIFs are calculated on 50 SNPs sliding windows over the entire chromosomes. With this threshold, 4,728,931 (81.9 %) of the total 5,772,170 SNPs were filtered out, and 1,043,239 (18.0%) were retained.

### eQTL Analysis

We computed the eQTL tests for the set of 18,072 protein coding genes for which we have sufficient coverage (see filter criteria for RNAseq data above). For each gene, we tested all SNPs in a 500kb cis window (500kb upstream from the TSS, 500kb downstream from the transcription termination site).

For each test, we model the effect of known clinical and technical covariates (Sex, Age, Batch, Smoking Status and PacksYear) using a fix effect. All clinical covariates were encoded as numerical values (0-4 for smoking status, 0-3 for age and packYears, and binary 0-1 for sex and batch), and genotypes are encoded as a numeric variable (0: Ref/Ref; 1: Alt/Ref; 2: Alt/Alt). P-values were computed using the R package Matrix eQTL (*49*). We used a two step multiple testing correction procedure, as described in (*50*). First, for each gene, we correct for the number of tests using Bonferroni correction. Second, we performed a global correction across the lead variants, that is, the most significant SNPs, per eQTL, using a Benjamini-Hochberg procedure.

### Gene environment interaction test

To test for a combined effect of genotype and environment on the gene expression level of the smoke injury gene, we conducted an interaction test between the genotype background and the smoking status of the patient, encoded in a 0/1/2 form (Never/Ex/Current). For each of the 749 smoke-injury gene, we retrieved the lead eQTL variant identified in the genome-wide eQTL analysis, and tested for an interaction effect between the genotype encoded in a 0/1/2 numeric and the smoking status, correcting for the effect of age, sex, smoking status, packyears, and genotype.

### Identification of GWAS-linked genes

To study the mechanisms by which germline genotype background influences the lung cancer risk, we adopted the approach developed by (*51*). We downloaded a curated set of 1261 GWAS lung cancer risk loci from the GWAS catalog (*52*) (see Sup. Tab. 9) and mapped genotyped and imputed SNPs of all patients to the nearest GWAS risk locus as follows. For each GWAS risk locus, we retrieved a list of variants in our cohort within a 500kb cis-window using a linkage disequilibrium (LD) cutoff of R^2>0.8 in the UK population using the Linkage Desiquilibrium Calculator of the ensembl website (*53*), yielding 9,739 candidate variants and 135,513 gene-SNP pairs. 3,455 of those 9,739 variants had a significant effect to their corresponding e-gene at a 5% FDR threshold. Many of those 3,230 hits were in LD with the same GWAS variant, such that all eQTL variants mapped to 67 unique GWAS risk loci (Sup. Tab. 8) from 10 different studies and were linked to the expression of 44 genes.

### Transcription factor network and activity

A context-specific protein-protein interaction network for nasal and bronchial epithelium was built using ARACNe-AP (*54*) on the vst-normalized expression data and a list of 1988 human transcriptional regulators, compiled using information available on public databases, from (*55*). ARACNe-AP was able to infer context-specific interactions across 1548 nasal and 1535 bronchial regulators. Activity of each of these regulators in each nasal and bronchial sample was inferred using *VIPER v1*.*20*.*0 (56)*. Network representations of TF-TF and TF-targets interactions were produced with *Cytoscape v3*.*8*.*1*. To find TFs that had an overrepresentation of GWAS genes in their target network, we used a context-specific TF-TF interaction network built using ARACNe-AP on bronchial vst-normalized gene expression data and a list of 1988 human transcriptional regulators (see above). For each TF *i*, we first counted the number (*N*_*G*_(*i*)) of genes in its target network that were identified as a GWAS gene. We then compared the proportion of GWAS genes in each TF target network to the expected number that would be found for a similar number of randomly selected genes with a one-tailed hypergeometric test using the phyper function in R with the following parameters:

**m**: total number of genes in the network of TF *i*; **n**: 18,062 m; **k**= the number identified of GWAS genes and **q** = *N*_*G*_(*i*), the number of GWAS genes in the target network of the TF *i*. Obtained p-values where adjusted for multiple testing using a Benjamini-Hochberg correction. We applied the same procedure to test for the enrichment of response genes in the 4 identified GWAS TFs, although we did not correct the p-values for multiple testing this time since we conducted only 4 tests.

## Data Availability

Data will be made available upon journal publication.

## Author contributions

MSDB and FMas processed and analysed the data, interpreted the results and wrote the manuscript; TTW, FG, MOR, IS and DS helped in data analysis and processing; RS contributed to experimental and study design; AS oversaw all sample and data collection; AG processed patient data and provided clinical classification; KM and FMar helped design and implement the study; RCR, BAJP and RFS designed, implemented and supervised the study, guided data analysis and wrote the manuscript; all authors agreed to the final version of the manuscript.

## Acknowledgements

RFS, MSDB and FMas would like to thank the Helmholtz Association for support. FMas was supported by a postdoctoral fellowship of the Fondation pour la Recherche Médicale (SPE201803005264). TTW was funded by the Deutsche Forschungsgemeinschar, CompCancer Research Training Group (RTG2424), project number 377984878. Computation has been performed on the HPC for Research cluster of the Berlin Institute of Health. Parts of this work were funded by CRUK core grant C14303/ A17197 and A19274 (FMar lab). This work was funded by grants to BAJP from Cancer Research UK and by the NIHR Cambridge Biomedical Research Centre (BRC-1215-20014). The views expressed are those of the authors and not necessarily those of the NIHR or the Department of Health and Social Care. BAJP is a Gibb Fellow of CRUK and NIHR Senior Investigator. RCR is part funded by the NIHR Cambridge Biomedical Research Centre (BRC-1215-20014), Cancer Research UK Cambridge Centre and Royal Papworth Hospital NHS Foundation Trust. Healthy volunteers were recruited through the Cambridge Bioresource (www.cambridgebioresource.group.cam.ac.uk). We thank the Bioinformatics and Genomics Core Facilities of the CRUK Cambridge Institute for their excellent support; and Prof. Paul Pharoah for advice. We thank Dr Doris Rassl and Radhika Prathalingham for advice about sample quality and processing. We thank the Royal Papworth Hospital Research and Development Department and Papworth Trials Unit Collaboration for overseeing the clinical phase of the work including their staff, Jenny Castedo, Theresa Green, Anne Joy, Tania Pettett, Victoria Senior, Anne Thomson and Victoria Tuck for assistance with sample and data collection. We thank Drs David Meek, Nick Carroll and Brendan Dougherty for help with bronchial sample collection. And a special thank you to Dr Lori Calvert for co-ordinating sample and data collection at Peterborough City Hospital.

## Competing Interests

FMar is founder, director and shareholder of Tailor Bio.

**Sup. Table 2:**
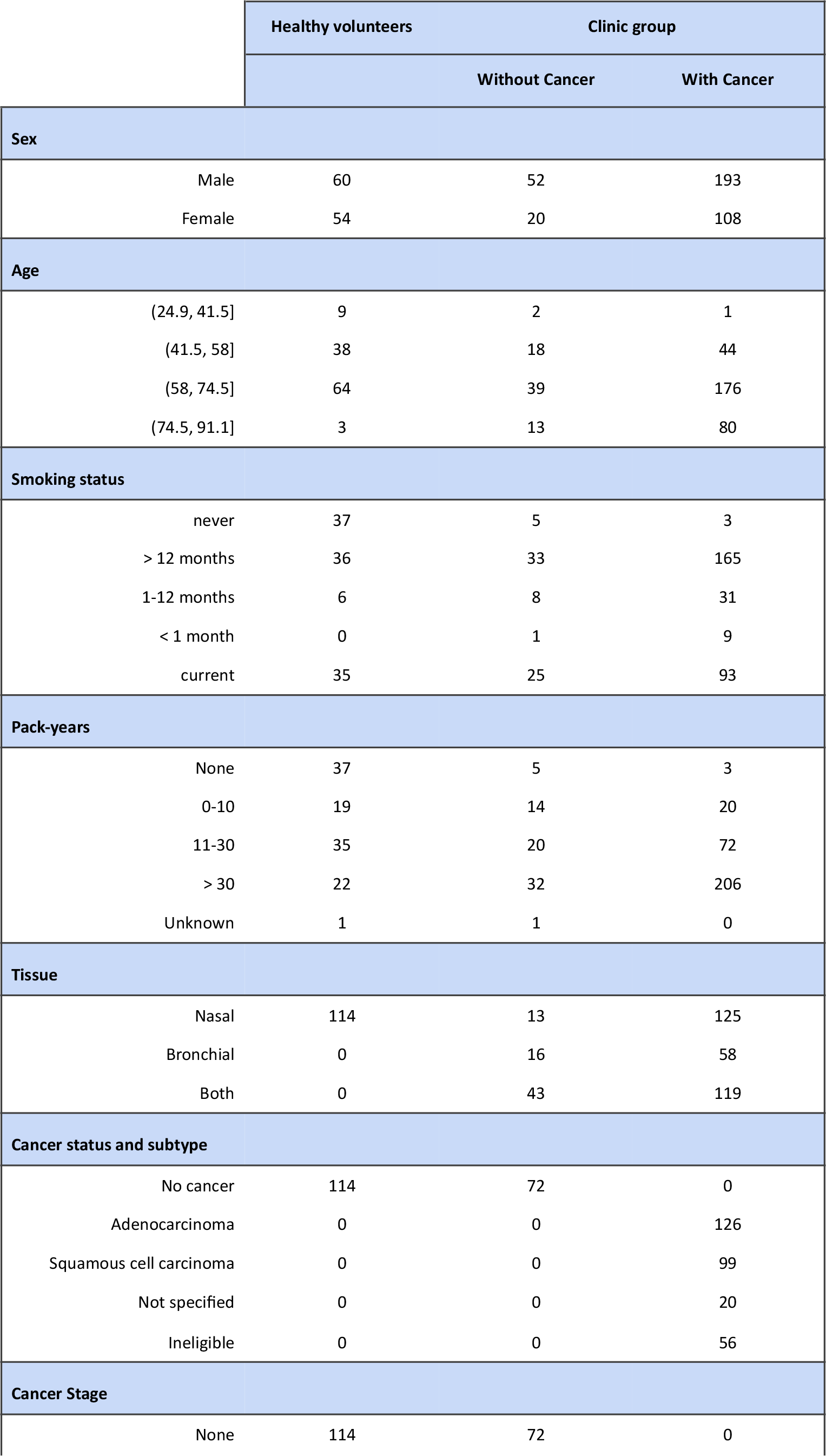
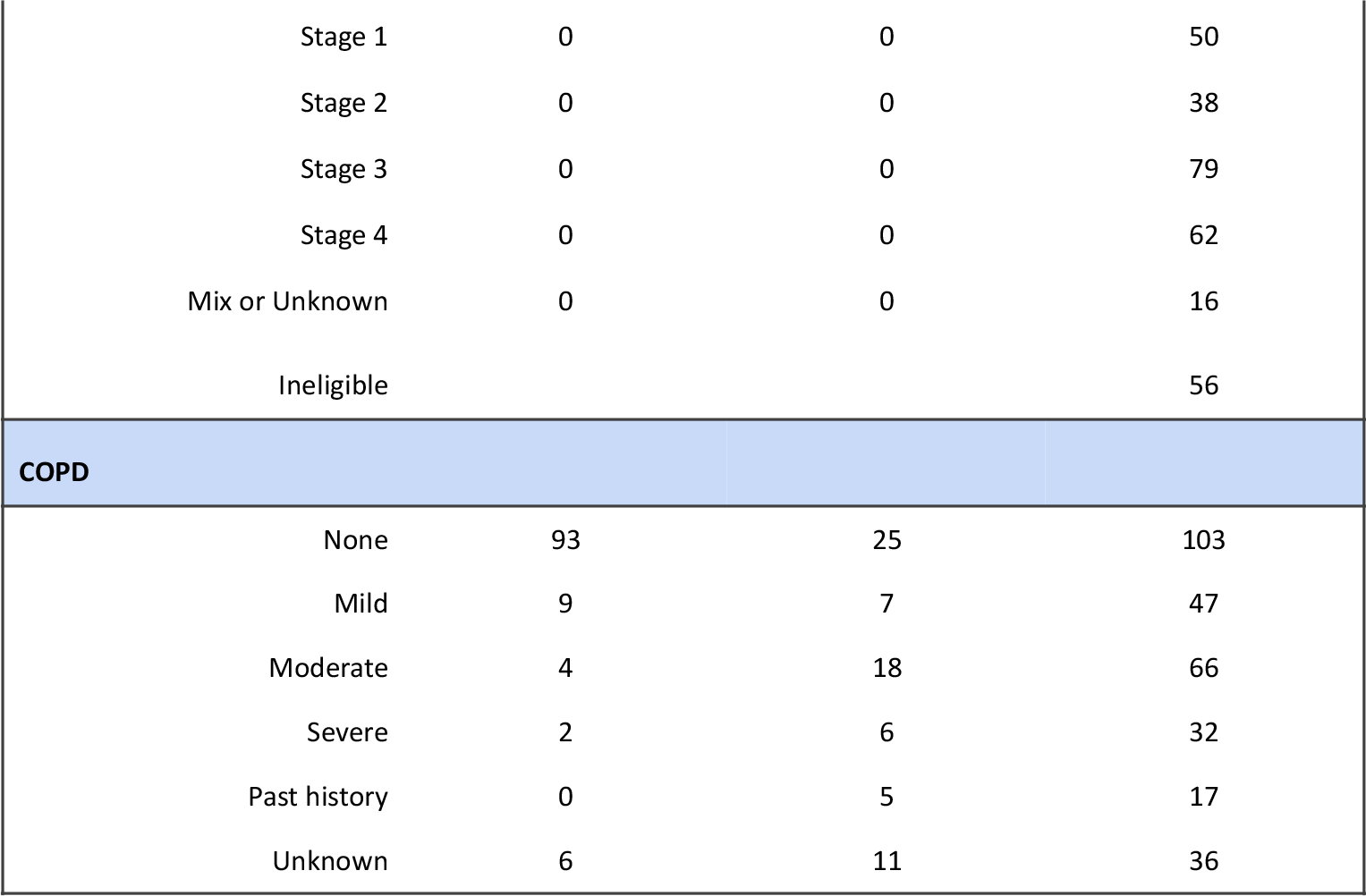
Clinical and demographic characteristics of the study subjects.

|  |  | Healthy volunteers | Clinic group |  |
| --- | --- | --- | --- | --- |
|  |  |  | Without Cancer | With Cancer |
| **Sex** |  |  |  |  |
|  | Male | 60 | 52 | 193 |
|  | Female | 54 | 20 | 108 |
| **Age** |  |  |  |  |
|  | (24.9, 41.5] | 9 | 2 | 1 |
|  | (41.5, 58] | 38 | 18 | 44 |
|  | (58, 74.5] | 64 | 39 | 176 |
|  | (74.5, 91.1] | 3 | 13 | 80 |
| **Smoking status** |  |  |  |  |
|  | never | 37 | 5 | 3 |
|  | > 12 months | 36 | 33 | 165 |
|  | 1-12 months | 6 | 8 | 31 |
|  | < 1 month | 0 | 1 | 9 |
|  | current | 35 | 25 | 93 |
| **Pack-years** |  |  |  |  |
|  | None | 37 | 5 | 3 |
|  | 0-10 | 19 | 14 | 20 |
|  | 11-30 | 35 | 20 | 72 |
|  | > 30 | 22 | 32 | 206 |
|  | Unknown | 1 | 1 | 0 |
| **Tissue** |  |  |  |  |
|  | Nasal | 114 | 13 | 125 |
|  | Bronchial | 0 | 16 | 58 |
|  | Both | 0 | 43 | 119 |
| **Cancer status and subtype** |  |  |  |  |
|  | No cancer | 114 | 72 | 0 |
|  | Adenocarcinoma | 0 | 0 | 126 |
|  | Squamous cell carcinoma | 0 | 0 | 99 |
|  | Not specified | 0 | 0 | 20 |
|  | Ineligible | 0 | 0 | 56 |
| **Cancer Stage** |  |  |  |  |
|  | None | 114 | 72 | 0 |

|  |  |  |  |
| --- | --- | --- | --- |
| Stage 1 | 0 | 0 | 50 |
| Stage 2 | 0 | 0 | 38 |
| Stage 3 | 0 | 0 | 79 |
| Stage 4 | 0 | 0 | 62 |
| Mix or Unknown | 0 | 0 | 16 |
| Ineligible |  |  | 56 |
| **COPD** |  |  |  |
| None | 93 | 25 | 103 |
| Mild | 9 | 7 | 47 |
| Moderate | 4 | 18 | 66 |
| Severe | 2 | 6 | 32 |
| Past history | 0 | 5 | 17 |
| Unknown | 6 | 11 | 36 |

## Notes

### Competing Interest Statement

FMar is founder, director and shareholder of Tailor Bio. All other authors declare no conflicts of interest.

### Funding Statement

FMas was supported by a postdoctoral fellowship of the Fondation pour la Recherche Medicale (SPE201803005264). TTW was funded by the Deutsche Forschungsgemeinschaft, CompCancer Research Training Group (RTG2424), project number 377984878. Parts of this work were funded by CRUK core grant C14303/A17197 and A19274 (FMar lab). This work was funded by grants to BAJP from Cancer Research UK and by the NIHR Cambridge Biomedical Research Centre (BRC-1215-20014). RCR is part funded by the NIHR Cambridge Biomedical Research Centre (BRC-1215-20014), Cancer Research UK Cambridge Centre and Royal Papworth Hospital NHS Foundation Trust.

### Author Declarations

Research ethics approvals for sample collection from participants in this study were given by East of England Cambridge Central REC 13/EE/0012 and the National Research Ethics Service Committee South East Coast - Surrey 13/LO/0889.

### Summary of Updates

Text shortened and made clearer; Updated figures; updated supplementary figures.

