## Supplementary Figures for "Smoking-dependent expression alterations in nasal epithelium reveal immune impairment linked to germline variation and lung cancer risk"

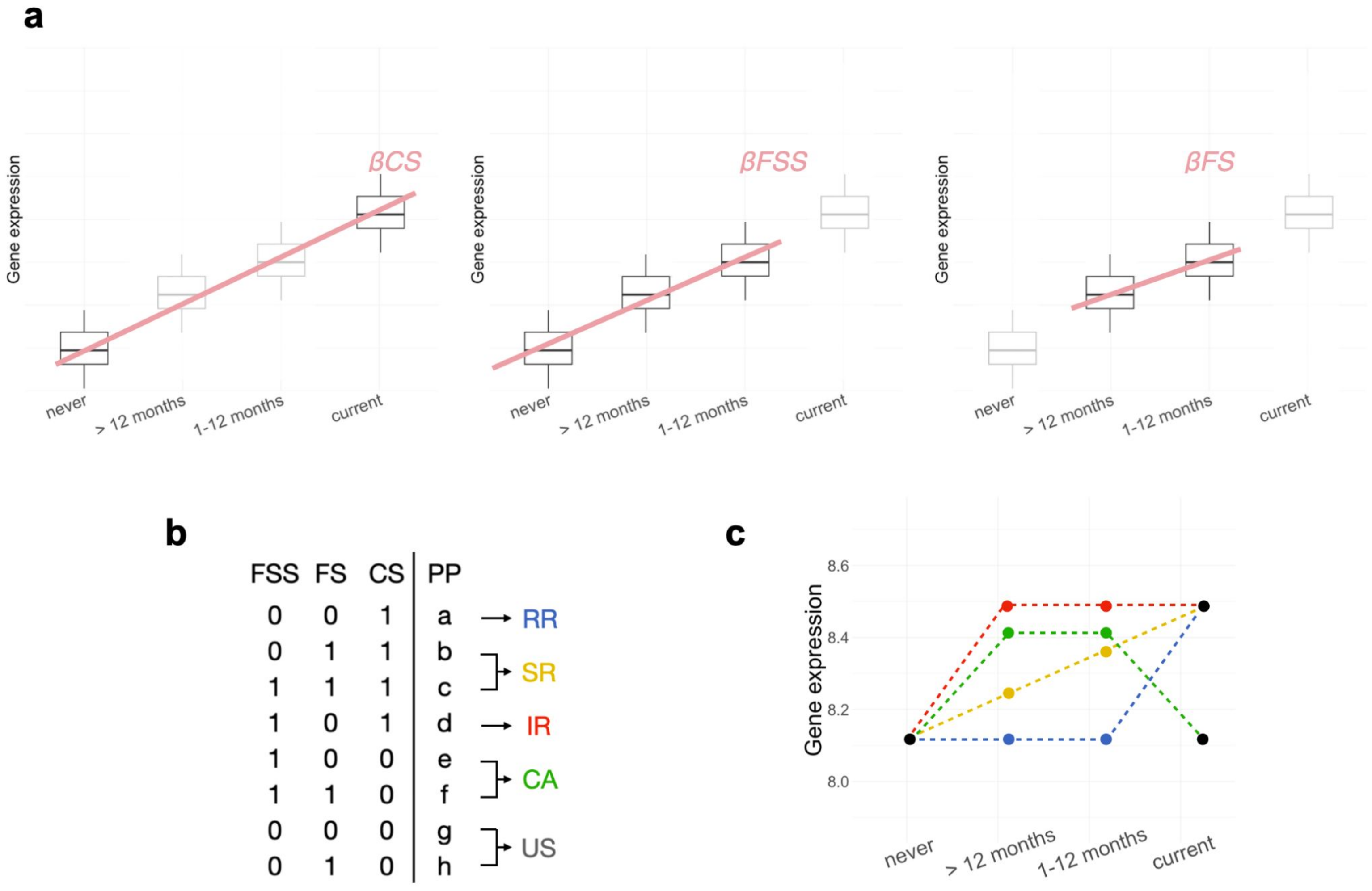


**Supp. Fig. 1 Smoke injury reversibility analysis.** (**a)** The slope coefficients associated to the three smoking status variables included in the Bayesian model (CS: current smoker status, FSS: former smoker status, FS: former smoker's time since quit). **(b)** Description of model selection procedure used to assign each gene to a reversibility class: the table shows all possible combinations of inclusion/exclusion of the three smoking status variables. **(c)** In blue, yellow and red, schematic of a gene with altered expression in current compared to never smokers, and the three possible trajectories after smoking cessation, corresponding to the RR, SR and IR reversibility classes; in green, schematic of a gene with no expression different in current versus never smokers, but altered expression in former smokers, corresponding to the CA class.
US: not affected; RR: rapidly reversible SR: slowly reversible; IR:irreversible; CA: cessation-associated.

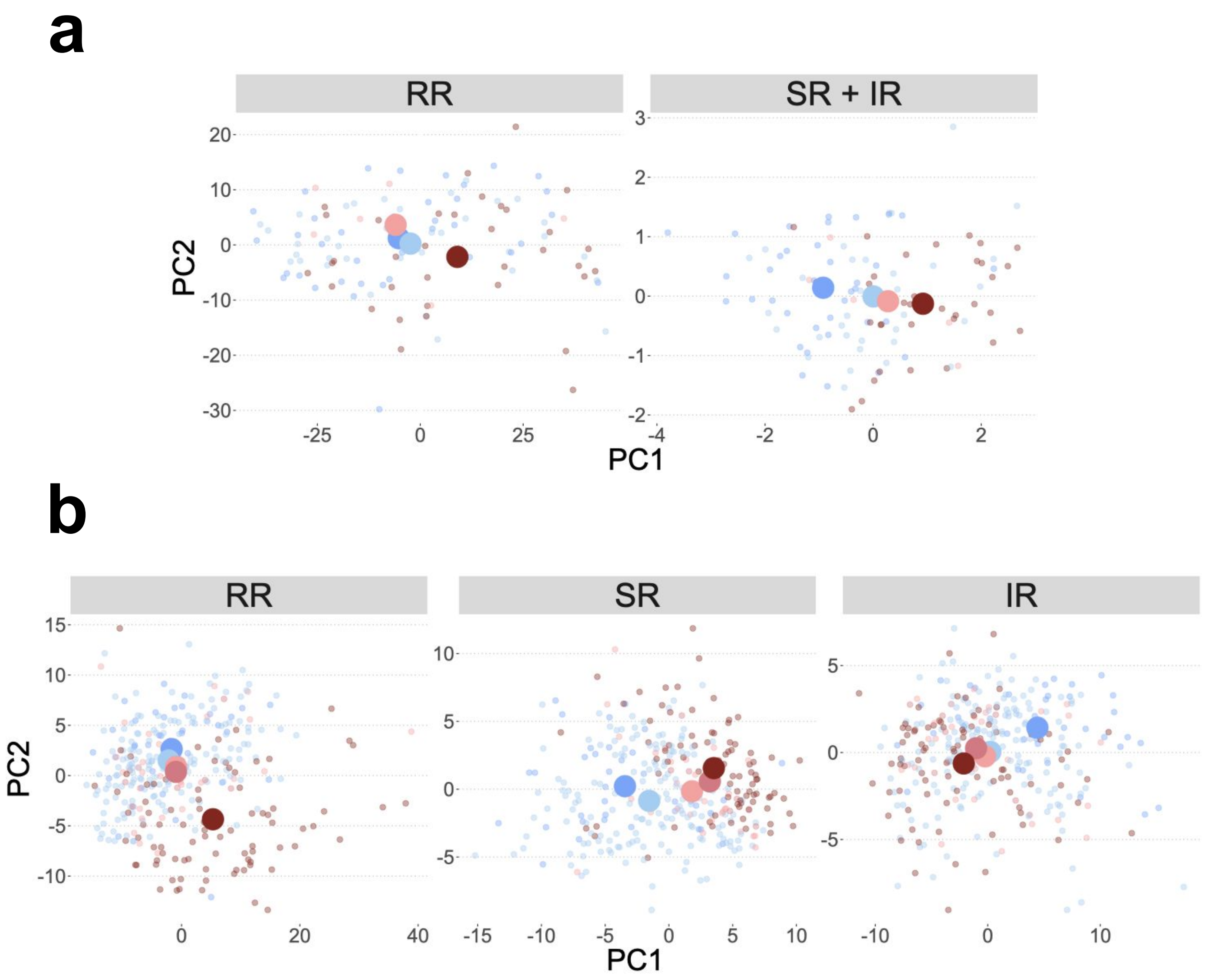


**Supp. Fig. 2 Principal component analysis on the genes belonging to different reversibility classes.** RR: Rapidly reversible genes, SR: Slowly reversible genes; IR:irreversible genes. Each small dot is a patient and colors indicate the smoking status of the patient. Large dots represent the mean of all patients for each smoking class. **(a):** nasal samples from healthy volunteers, using the reversibility classes from the bayesian model on the healthy volunteer group. Since only 2 genes were classified as IR, PCA was performed jointly for SR and IR genes **(b):** nasal samples from clinic subjects (cancer + benign), using the reversibility classes from the bayesian model on the clinic group.

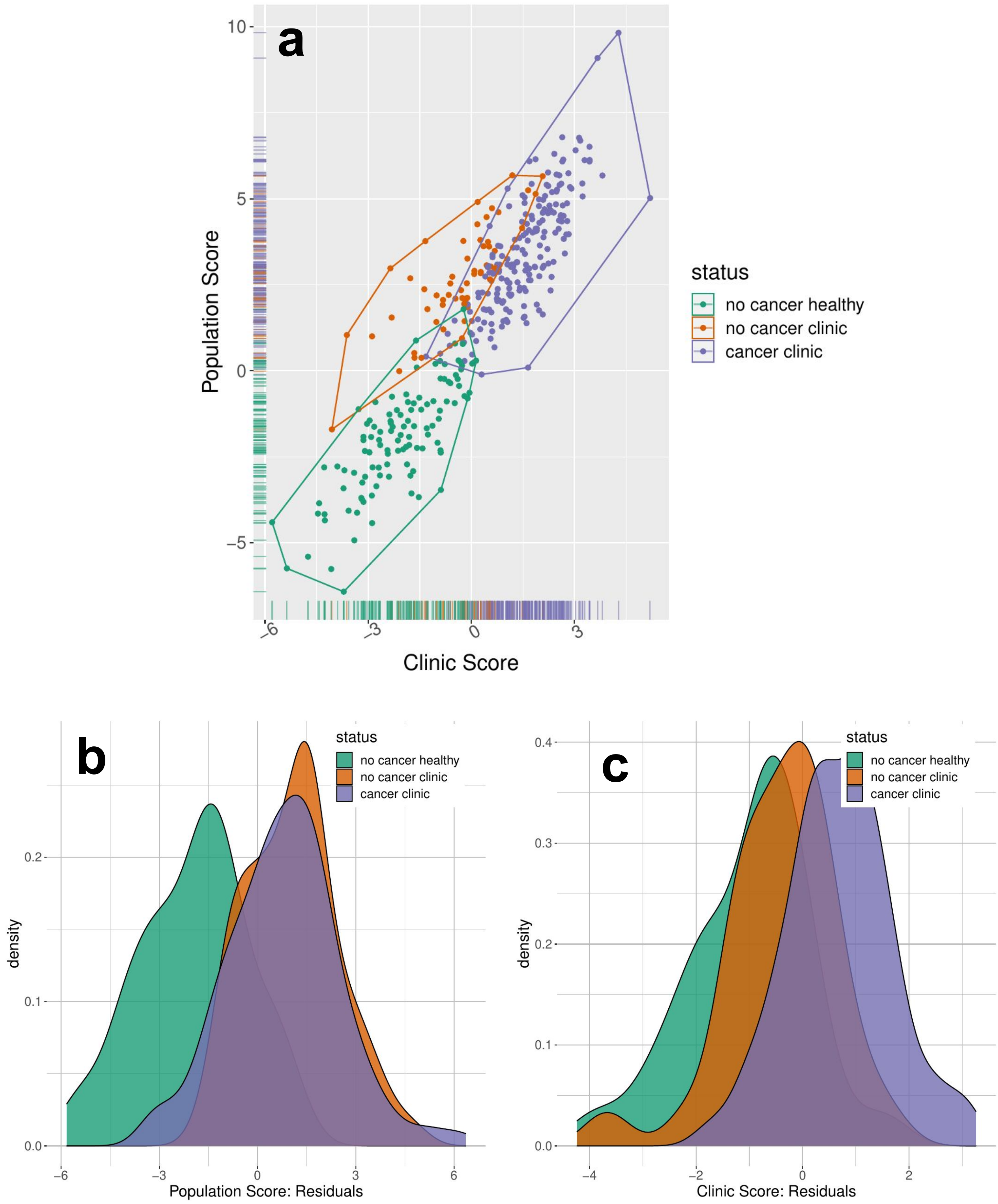


**Supp. Fig. 3: Population and clinic risk scores. (a)** Correlation between the clinic risk score and the population risk score for each patient. Each dot represent a single patient (green: healthy volunteer; orange: clinic benign; purple: clinic cancer). **(b-c)**: Distribution of the risk scores after regressing the for the population risk score **(b)** and the clinic risk score **(c)**.

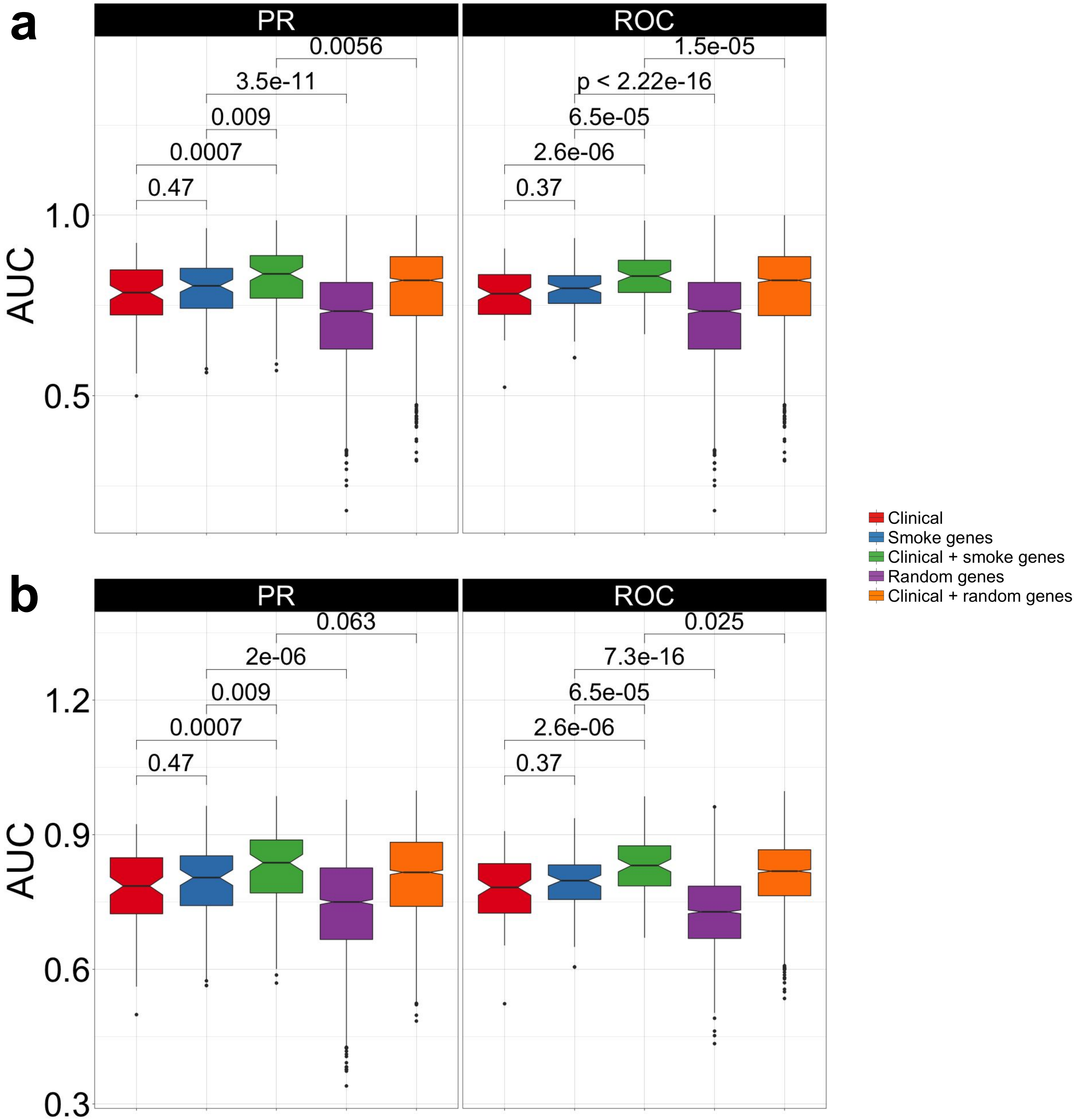


**Supp. Fig. 4: Area under the Curve (AUC) calculated after cross validation for different models.** Clinical: model trained on clinical data only, Smoke genes: model trained on the expression of smoke injury risk genes only, Clinical + smoke genes: model trained on clinical data and expression of smoke injury risk genes, Random genes: model trained on expression of a set of randomly selected genes, Clinical + random genes: model trained on clinical data and expression of a set of randomly selected genes. **(a)** Area under the PR and ROC curve for the population score, **(b)** Area under the PR and ROC curve for the clinic score.

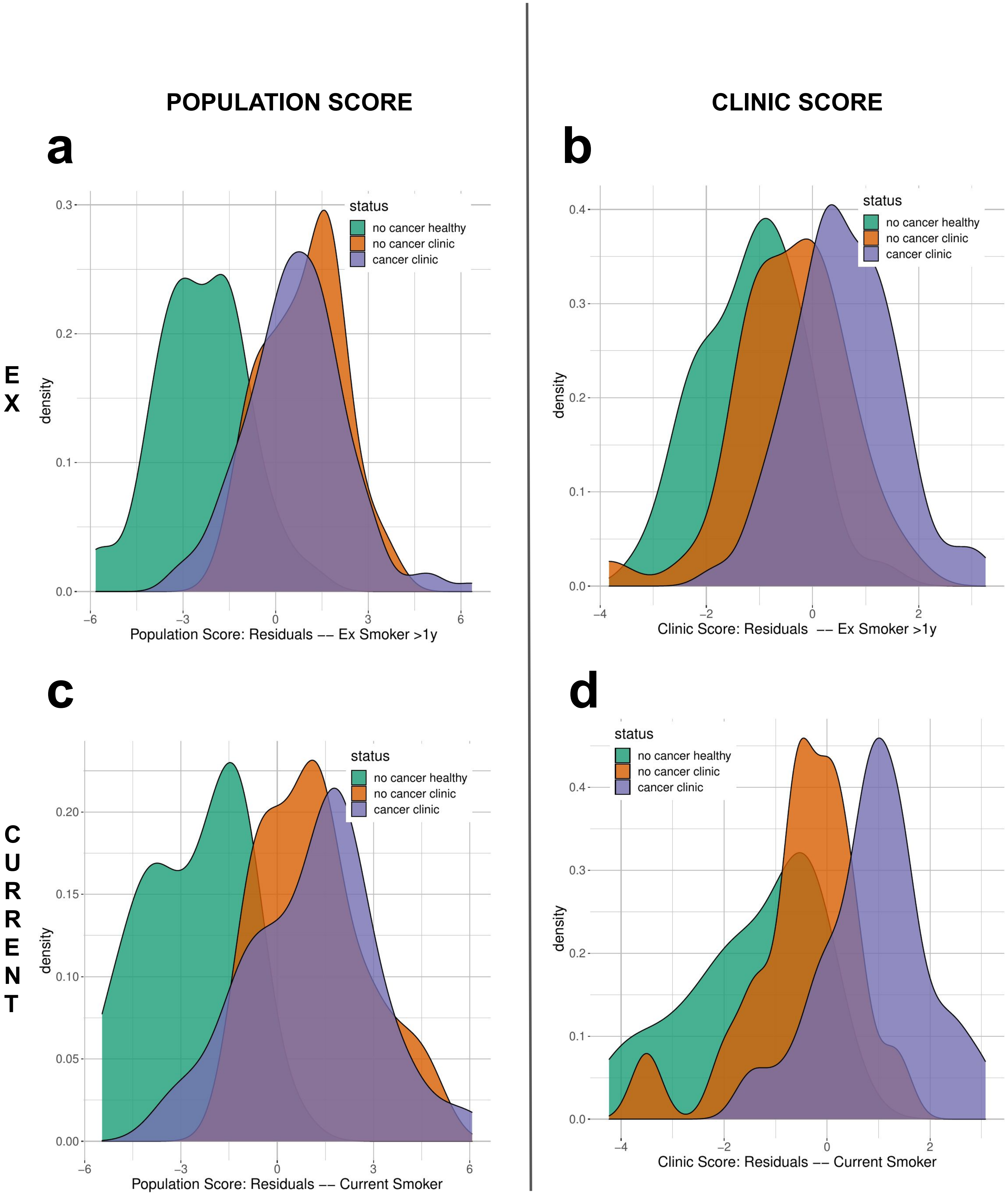


**Supp. Fig. 5: Risk scores stratified by smoking status**. Distribution of the population and clinic risk scores in ex smokers who stopped smoking for more than 1 year (**a-b**) and in current smokers (**c-d**), after regressing out clinical covariates.

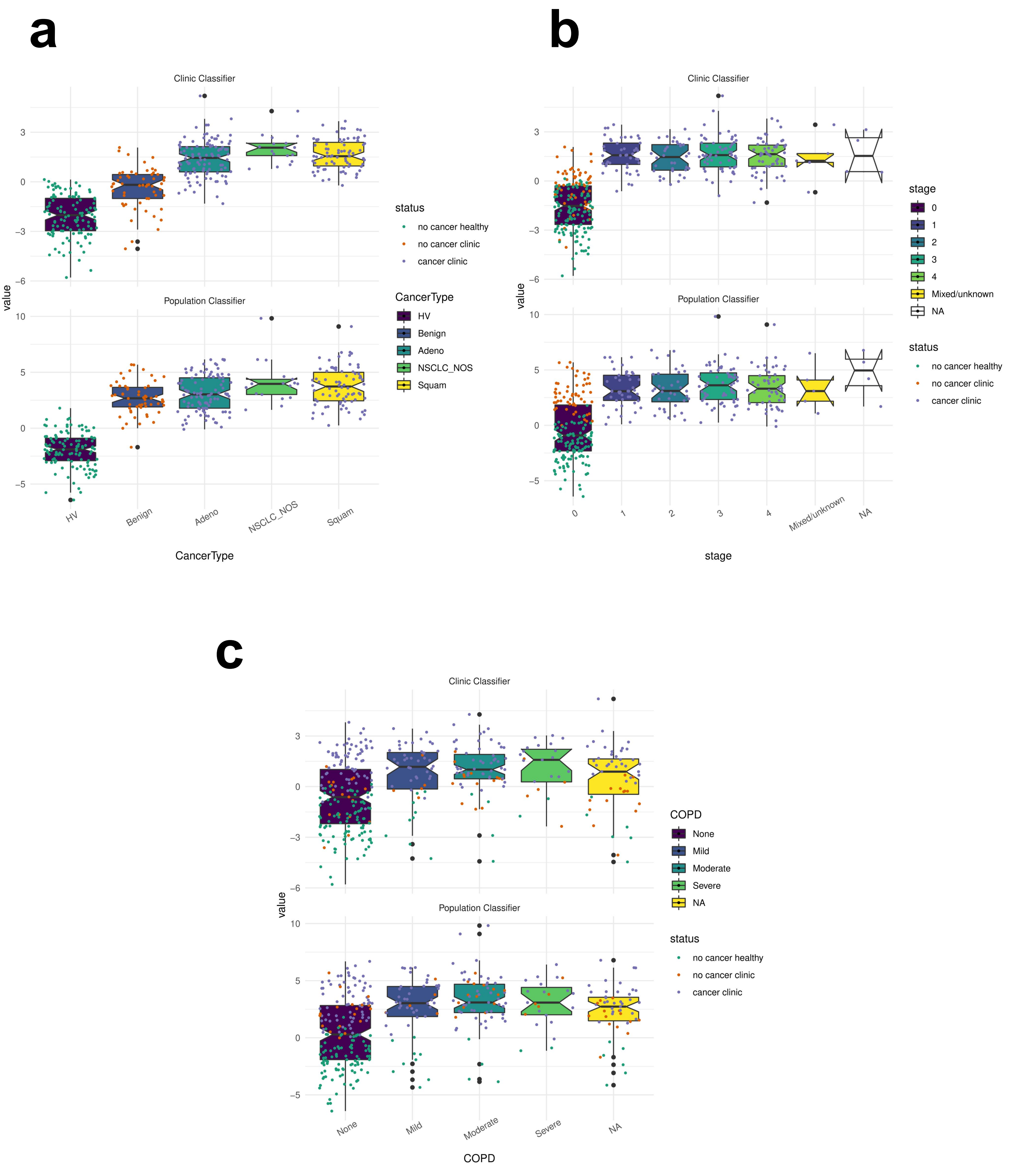


**Supp. Fig. 6: Clinic and population scores for different clinical variables**. Distribution of the clinic (**Top rows)** and population **(bottom rows)** risk score in subjects depending on (**a**) The type of cancer (**b**) the stage of the cancer (**c**) the COPD status. Color of the dot indicate for each individual subject his status, namely healthy volunteer (green), clinic benign (orange) or clinic cancer (purple)

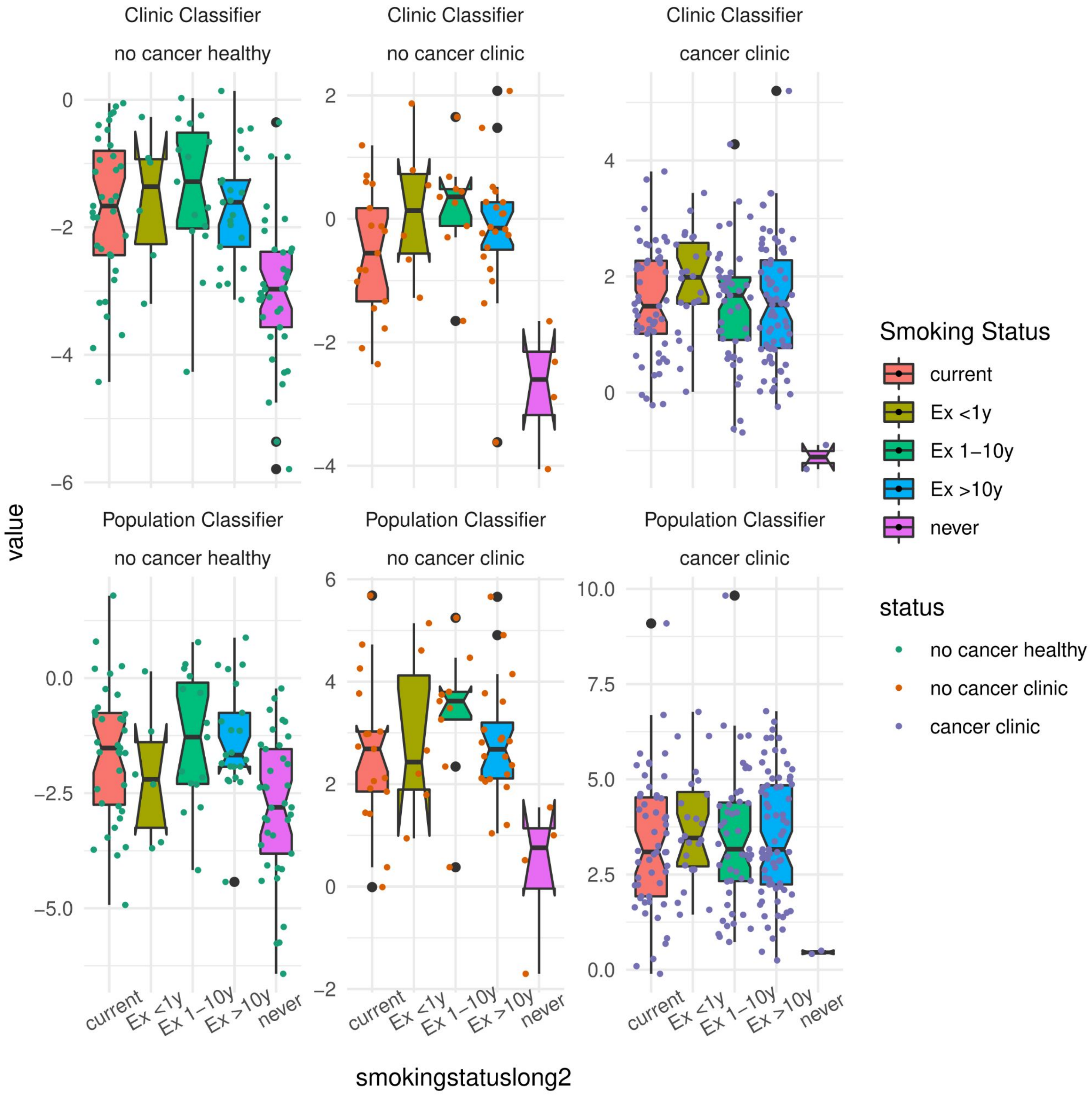


**Supp. Fig. 7: Clinic and population scores depending on smoking status**: Distribution of the clinic (**Top row)** and population **(bottom row)** risk score in subjects depending on their smoking status. Scores are represented separately for healthy volunteers (left), clinic benign (middle) and clinic cancer (right). Color of the dot indicate for each individual subject their status, namely healthy volunteer (green), clinic benign (orange) or clinic cancer (purple)

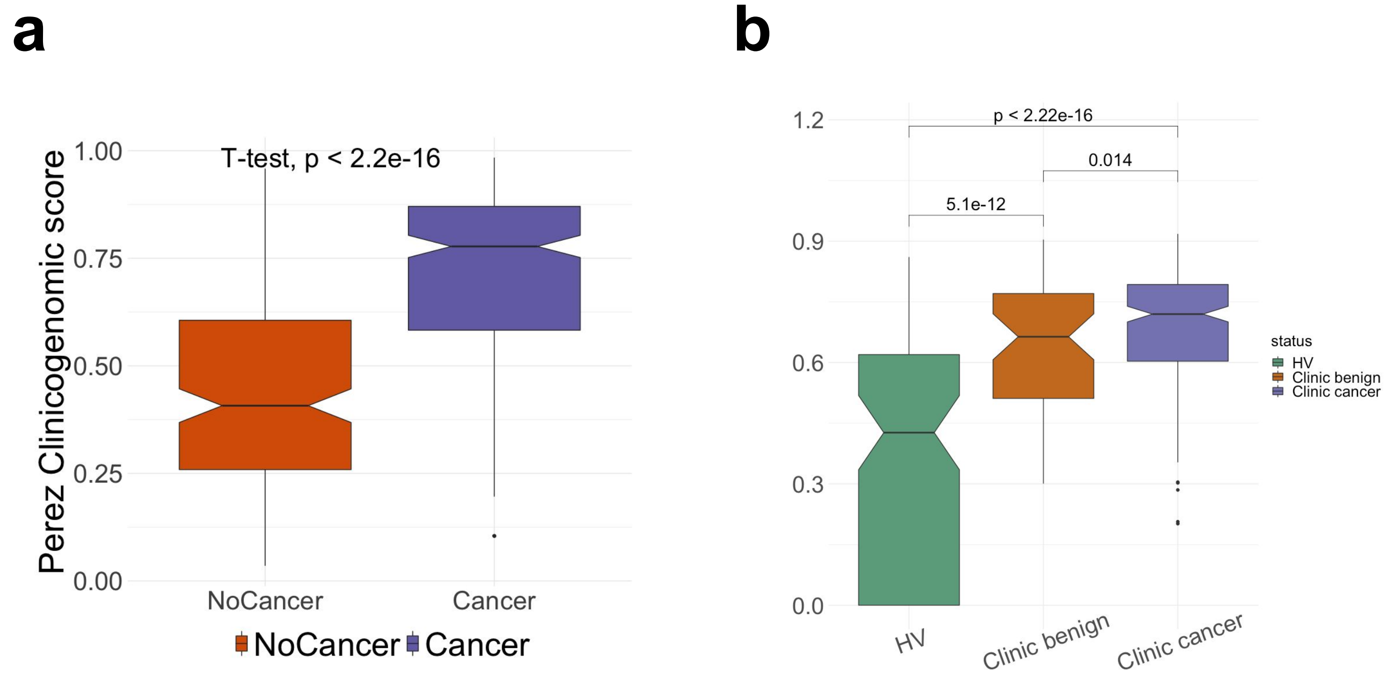


**Supp. Fig. 8: Comparison with classifier from Perez-Rogers et al. (2017). (a)** Perez-Rogers' clinico-genomic model applied to the AEGIS nasal cohort; **(b)** Perez-Rogers' clinico-genomic model applied to nasal samples from our cohort.

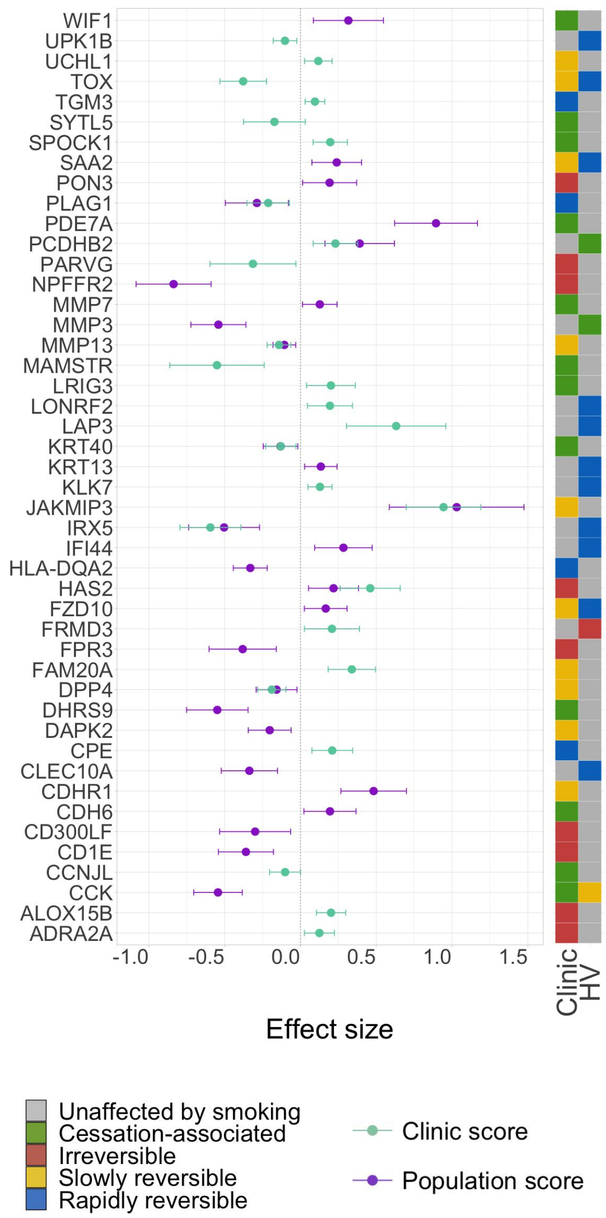


**Supp. Fig. 9: Genes robustly contributing to the population and clinic risk scores.** The weight of the genes selected in more than 80% of cross validations in the population and clinic classifiers; the presented value is the mean over all cross validation and the error bars represent standard deviation; the annotation track on the right shows the reversibility classes of the genes in the HV and clinic groups.

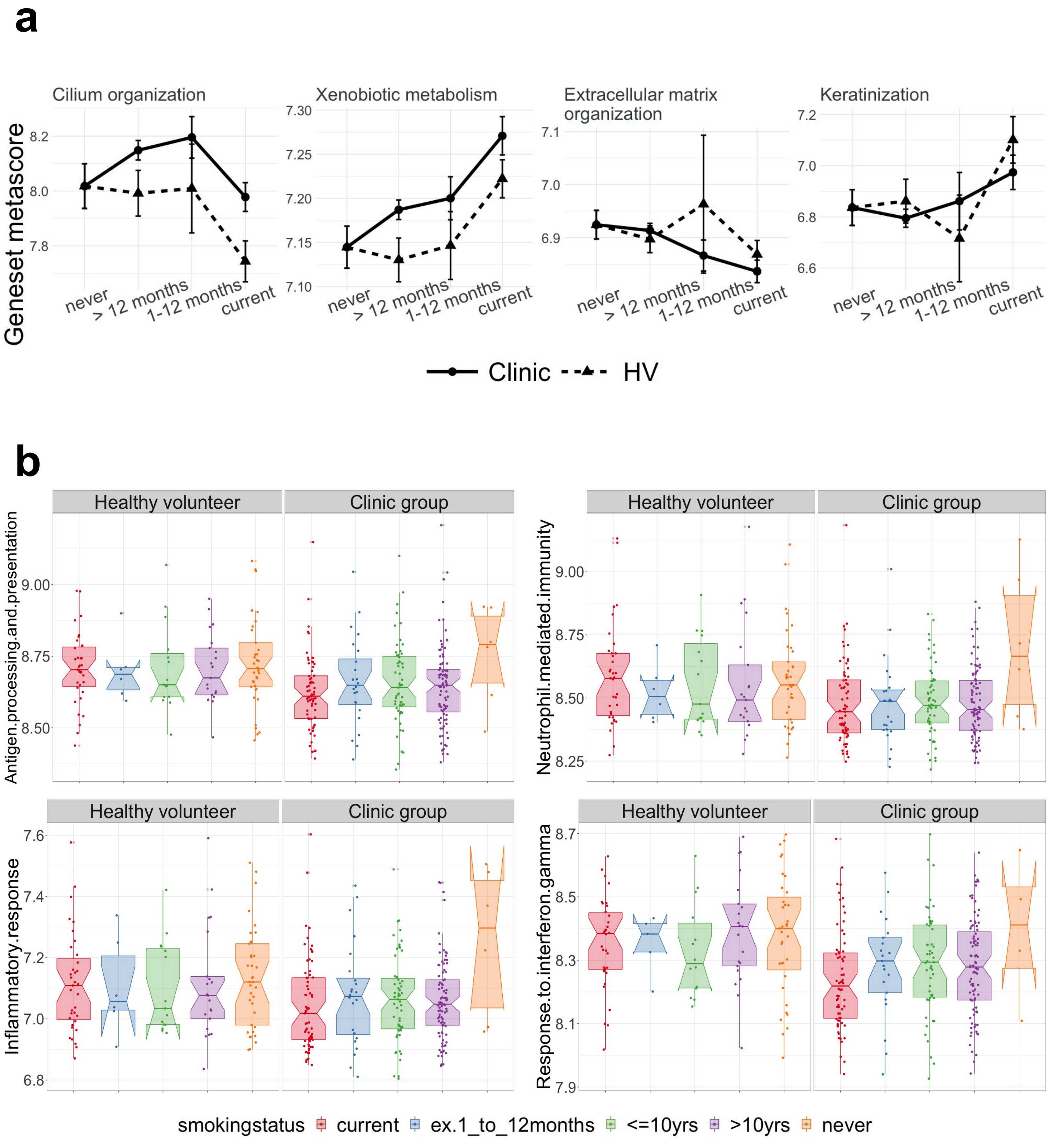


**Supp. Fig. 10: Pathway metascore over smoking status. (a)** Four GO terms involved in smoke injury response in nasal epithelium; **(b)** Immune-related genesets; former smokers with time since quit > 1 year were divided into former smokers who quit <= 10 years and >10 years before sample collection. Geneset metascores were calculated by averaging the nasal expression of genes belonging to each GO term.

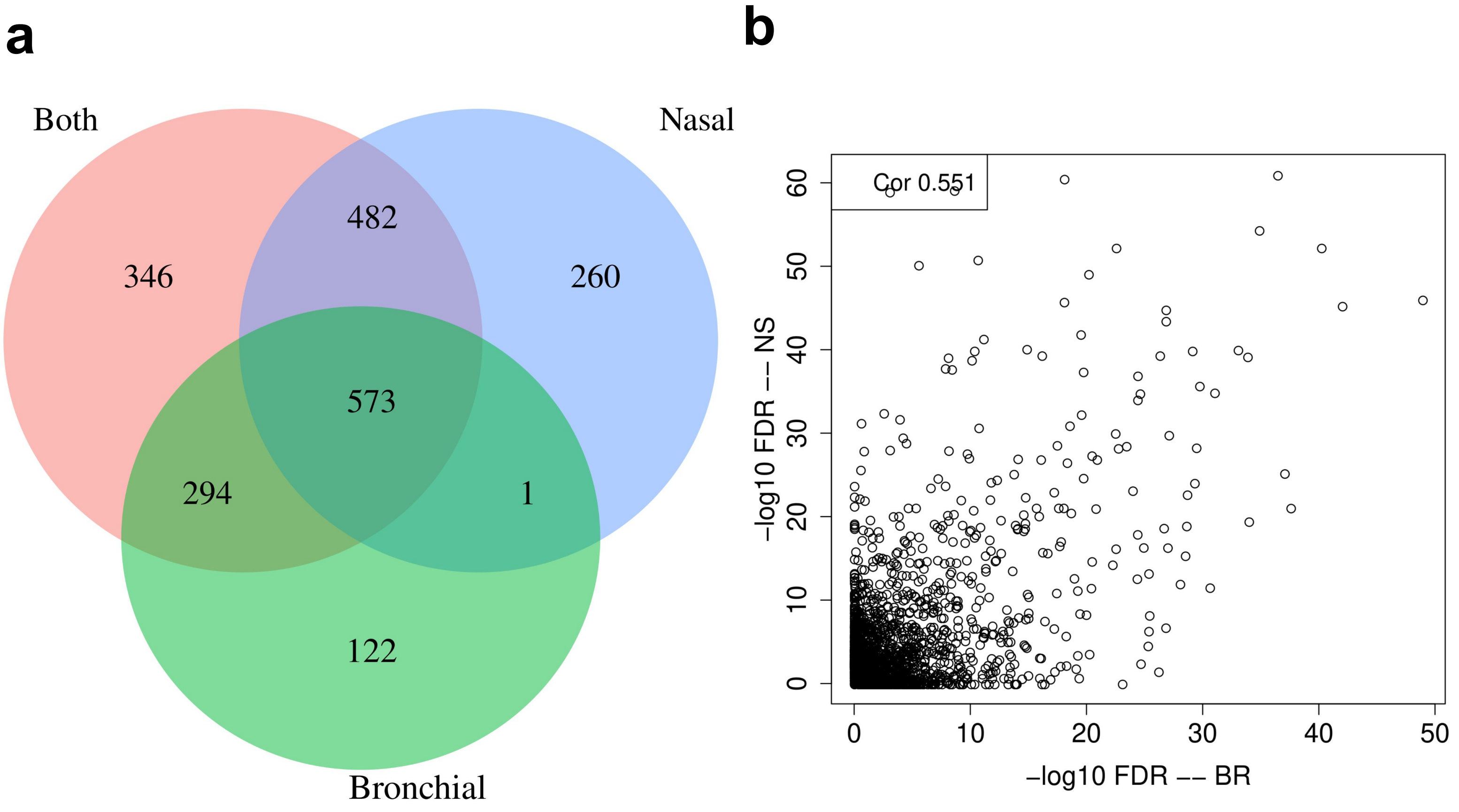


**Supp. Fig. 11: Overlap of eQTL Analysis.** eQTLs in the nasal and bronchial tissue are strongly correlated. **a:** A venn diagram of the eQTL genes found in the nasal tissue (blue), in the bronchial tissues (green) or in an analysis conducted jointly in the two tissued (red). **b:** Correlation between the corrected p-values (-log10) of the eQTL test in all tested genes between the test conducted in the bronchial (BR) and nasal (NS) tissues.

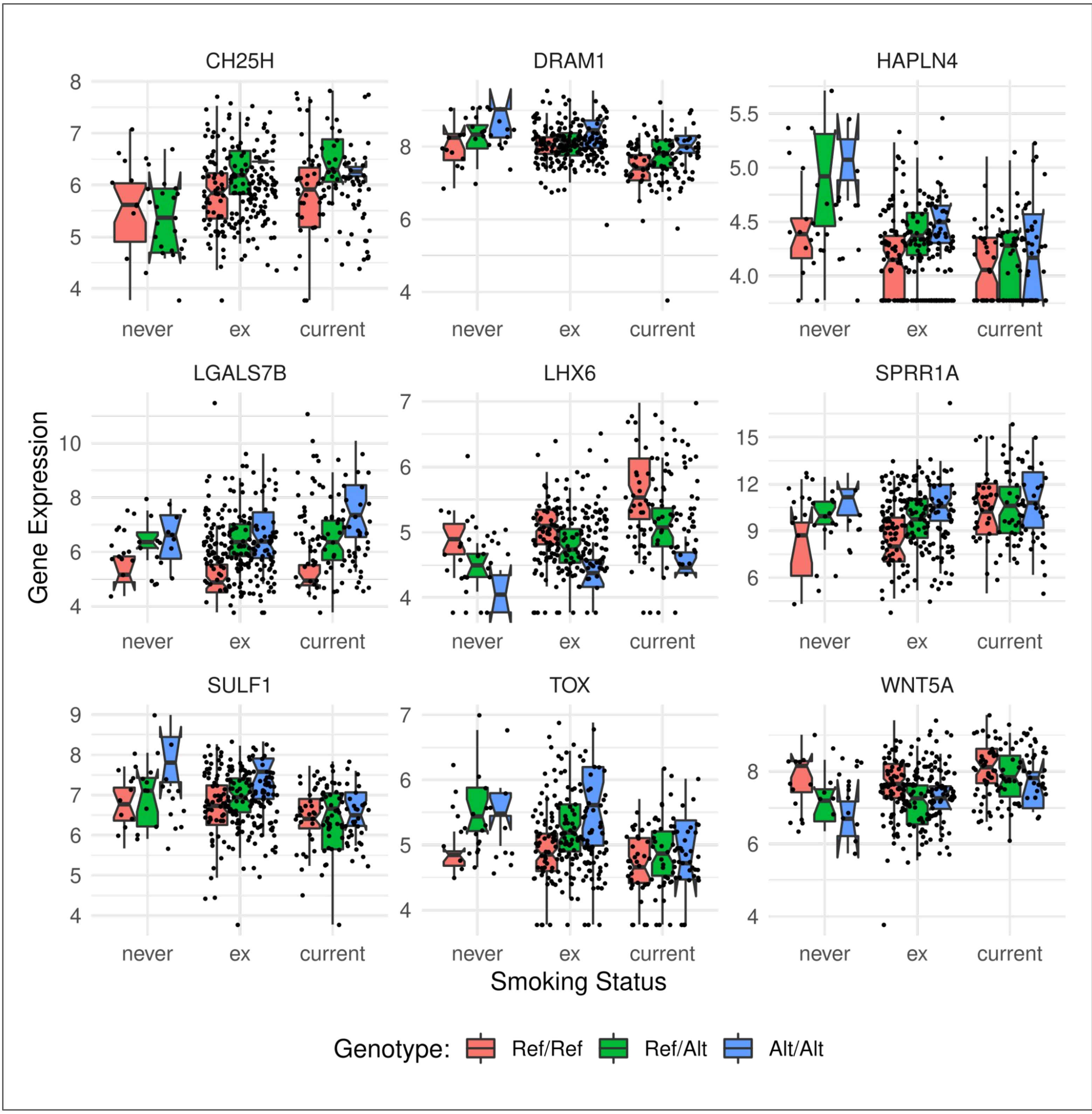


**Supp. Fig. 12: Combined environmental and genetic effect on the expression for 9 gene in nasal tissues.** For the 9 genes with a significant interaction effect between smoking status and the genotype of the patient at the lead eQTL position, we present the expression level of the gene separately for never, former and current smokers. Samples are further stratified depending on the genotype of the subject at the corresponding lead eQTL locus (pink: homozygous reference; green: heterozygous; blue homozygous Alternative). P-values and SNP position are given in Supp Table 6.

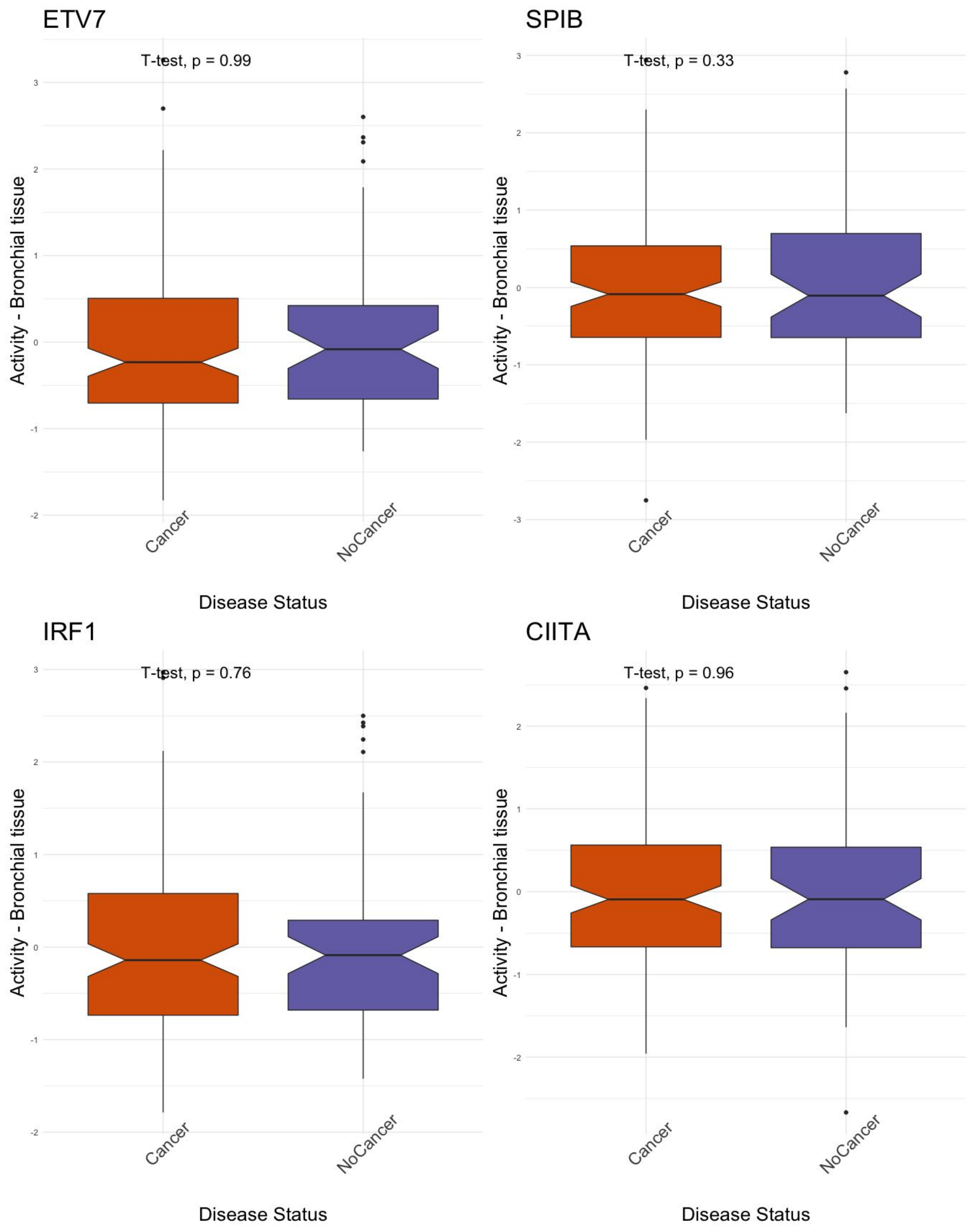


**Supp. Fig. 13**: Equivalent to Fig 6b, the activity level of the 4 TFs that regulate a high number of risk and GWAS genes, but this time calculated for the Bronchial samples only, on a gene network that is inferred from the Bronchial samples. As in the nasal samples, we found no differences between clinic patients with and without cancer. We did not obtain bronchial tissue from healthy volunteers for ethical reasons.

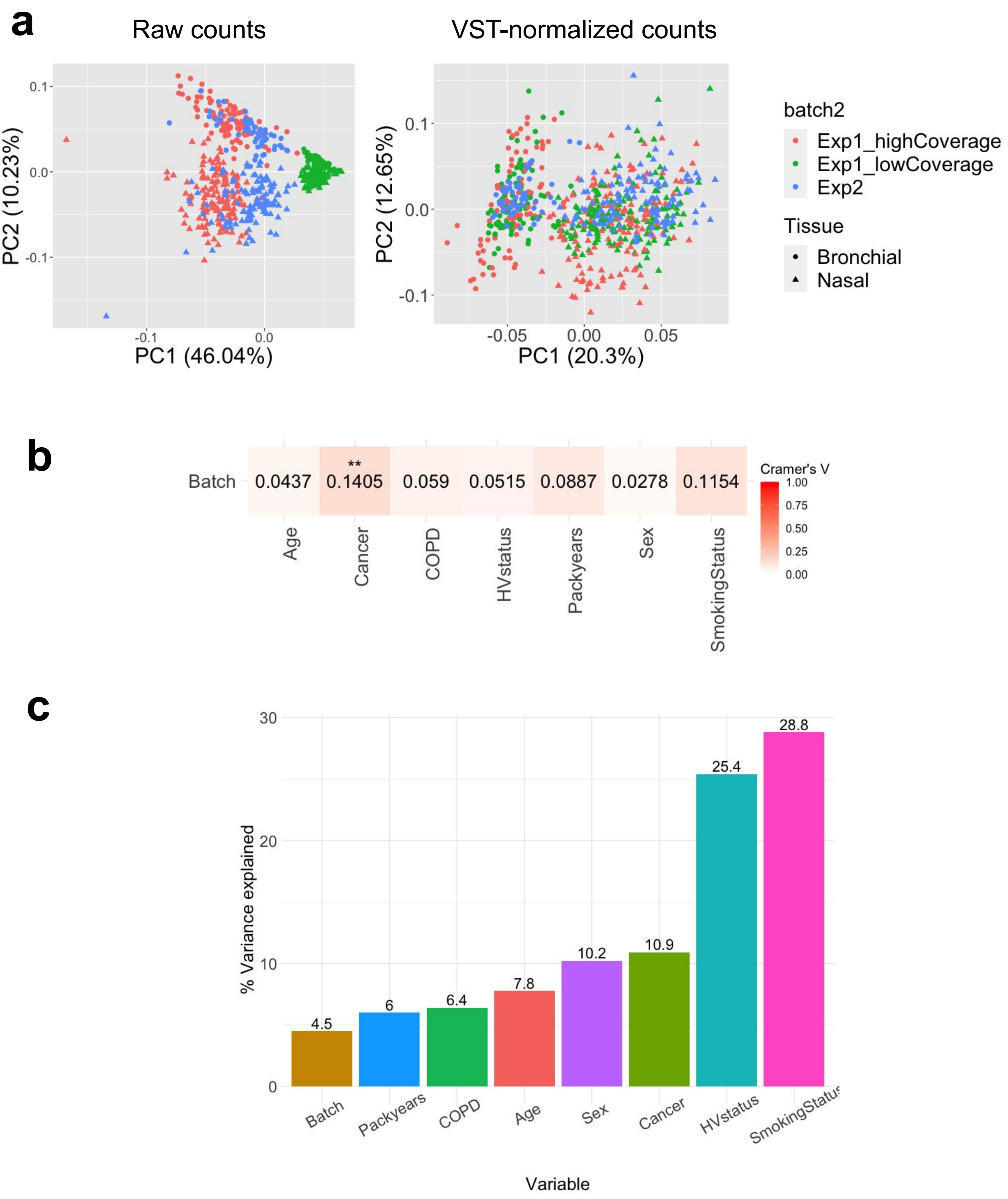


**Supp. Fig. 14 Exploratory analysis**. (**a)** PCA of all samples before (left) and after (right) VST-normalization. **(b)** Strength and significance of association between experimental batch and clinical covariates; for each pair of covariates Cramer's V value and chi-square test pvalue are reported (*: P <= 0.05, **: P <= 0.01, ***: P <= 0.001). (c) Contribution of different clinical variables to the total explained variance in gene expression calculated using a random model on nasal samples.
